# Early dynamics of circulating tumor DNA predict chemotherapy responses for patients with esophageal cancer

**DOI:** 10.1101/2021.02.25.21251979

**Authors:** Ryosuke Fujisawa, Takeshi Iwaya, Fumitaka Endo, Masashi Idogawa, Noriyuki Sasaki, Hayato Hiraki, Shoichiro Tange, Tomomi Hirano, Yuka Koizumi, Masakazu Abe, Tomoko Takahashi, Mizunori Yaegashi, Yuji Akiyama, Mari Masuda, Akira Sasaki, Fumiaki Takahashi, Yasushi Sasaki, Takashi Tokino, Satoshi S. Nishizuka

## Abstract

**Purpose:** We investigated whether early circulating tumor DNA (ctDNA) changes, measured using digital PCR (dPCR), can predict later chemotherapy responses in esophageal squamous cell cancer (ESCC).

**Design:** We compared the dynamics of ctDNA and tumor volumes during chemotherapy in 42 ESCC. The accuracy of predictions of later chemotherapy responses were evaluated by the ratio of the variant allele frequency (VAF) of ctDNA (post-/pre-ctDNA) and the total tumor volume (post-/pre-volume) before and after an initial chemotherapy cycle using a receiver-operating characteristic curve analysis. Total positive and negative objective responses (ORs) were defined as either >50% or ≤50% reductions, respectively, in the total tumor volume at the end of first-line chemotherapy.

**Results:** Mutation screening of 43 tumors from 42 patients revealed 96 mutations. The pretreatment dPCR-ctDNA data were informative in 38 patients, using 70 selected mutations (1–3 per patient). The areas under the curve (AUCs) for the post-/pre-volume and post-/pre-ctDNA levels used in predicting the total OR were 0.85 and 0.88, respectively. The optimal cutoff value of post-/pre-ctDNA was 0.13. In 90% (18/20) of patients with a post-/pre-volume ≥50%, the total OR could be predicted by the post-/pre-ctDNA with high accuracy; the AUC by post-/pre-ctDNA was higher than that by post-/pre-volume (0.85 vs 0.76, respectively). Patients with low post-/pre-ctDNA (n = 18) had a significantly better overall survival rate than those with high post-/pre-ctDNA (n = 20; *P* = 0.03).

**Conclusions:** Early ctDNA changes after an initial cycle of chemotherapy predict later responses to treatment with high accuracy in ESCC patients.

## Introduction

Esophageal squamous cell cancer (ESCC) is a malignant tumor with a poor prognosis; the 5-year survival rate is <20% (1). Systemic chemotherapies have been playing an important role in the treatment of advanced ESCC. For example, combination therapy with cisplatin and fluorouracil (CF) has been in use since the 1990s (2,3). Because locally advanced or metastatic ESCC’s response rate to CF has been reported as below 40%, neoadjuvant setting of CF for resectable ESCC and alternative regimens of CF plus another drug have been introduced to improve survival outcomes (2). Neoadjuvant CF prior to surgery improved survival rates, and has been regarded as a standard regimen for Stage II/III ESCC patients (4). CF plus docetaxel (DCF) demonstrated a high response rate of about 60% (5,6). Many chemotherapy regimens for ESCC have been reported, with most treatments given in repeating cycles and with combination chemotherapy cycles usually taking 3-4 weeks each. Therefore, each regimen is usually continued for several months.

The current gold standard for assessing treatment response is the image-based Response Evaluation Criteria in Solid Tumors (RECIST) (7). However, it often has poor interobserver reproducibility, and a minority of patients are classified as not having “measurable disease.” Furthermore, the evaluation usually occurs 2 months after treatment, when a response will be confirmed. In ESCC patients, an “unmeasurable” primary tumor is often the largest lesion, and it is difficult to distinguish benign reactive enlargement from frequently occurring metastasized lymph nodes. In addition, the response to treatment often becomes evident after several cycles of chemotherapy. About 40–60% of ESCC patients treated with CF and DCF do not respond to chemotherapy. Until progressive disease is confirmed by imaging, the regimen is usually continued after each cycle of chemotherapy in patients evaluated as having stable disease (SD). Therefore, it is possible that about half of advanced ESCC patients receive several cycles of ineffective therapy. The early and accurate confirmation of chemotherapy’s efficacy is crucial to reduce unnecessary adverse effects and to switch to an alternate therapy promptly.

Analyses of circulating tumor DNA (ctDNA) levels have demonstrated promising results for predictions of relapses and evaluations of treatment efficacy in patients with several cancer types (8-12). Several reports also demonstrated that early ctDNA dynamics could predict later chemotherapy responses, progression-free survival rates, and overall-survival (OS) rates in some cancer types (10,13-17). However, the methodology used to evaluate ctDNA changes and tumor responses varied by tumor type.

In terms of ESCC, several reports have demonstrated that ctDNA analyses could provide information for accurate diagnoses of treatment effects and accurate predictions of both tumor recurrence/progression and risks of recurrence and death (18-20). These reports used next-generation sequencing (NGS) of plasma DNA to target from 53 to 607 genes. As NGS requires costly multiple samples/library preparations and analytical processes, the frequency and interval of ctDNA testing in each patient were limited, not only in ESCC but also other malignancies. Furthermore, as mentioned above, it was difficult to measure most ESCC tumor volumes accurately. Therefore, the association between early ctDNA dynamics and tumor responses has not been investigated in ESCC patients.

We previously demonstrated that digital PCR (dPCR) allows for frequent monitoring of the tumor burden using a small number of tumor-specific ctDNA, enabling predictions of relapses and treatment efficacy, as well as relapse-free corroborating in management of ESCC patients (21). In the study, ctDNA could trace both the emergence of small recurrent tumors and regression of the lesions after treatment for a relapse in a timely manner. This suggested that early ctDNA dynamics during chemotherapy reflect changes in tumor volumes more accurately than image diagnosis, and could predict what responses would be after several cycles of chemotherapy. In this study, we investigated whether detecting tumor-specific ctDNA changes by dPCR before and after an initial cycle of chemotherapy can predict later responses after repeating chemotherapy cycles.

## Materials and Methods

### Patients and sample collection

This study was approved by the Institutional Review Board of Iwate Medical University (IRB# HGH27-16 and HG2020-021). Written informed consent was obtained from all patients. Out of 64 patients enrolled in the study “Tumor burden monitoring with circulating tumor DNA in esophageal squamous cell carcinoma patients” (UMIN Clinical Trial Registry: UMIN000038724), 47 patients received chemotherapy as the first-line therapy. Of these, 5 patients did not have a blood sample taken after the initial cycle of chemotherapy. Consequently, 42 patients were enrolled in this study (**Fig. 1**). All patients were histologically confirmed to have ESCC and were registered between September 25, 2015, and July 19, 2019. The patient characteristics are listed in Supplementary Table S1. Endoscopically acquired pretreatment primary tumor tissue samples and corresponding serial blood samples were obtained from all enrolled patients. In principle, the imaging examinations and blood collection for ctDNA and serum tumor markers (TM), including squamous cell carcinoma antigens (SCC), carcinoembryonic antigens (CEA), and cytokeratin 19 fragments (CYFRA 21-1), were completed at the same time. During chemotherapy, treatment efficacy was evaluated after each 3-to 5-week cycle. DNA was extracted from tumor tissues, peripheral blood mononuclear cells (PBMC), and a series of plasma samples. The methodologies used to extract DNA from each sample type were described previously (21).

**Figure 1.**
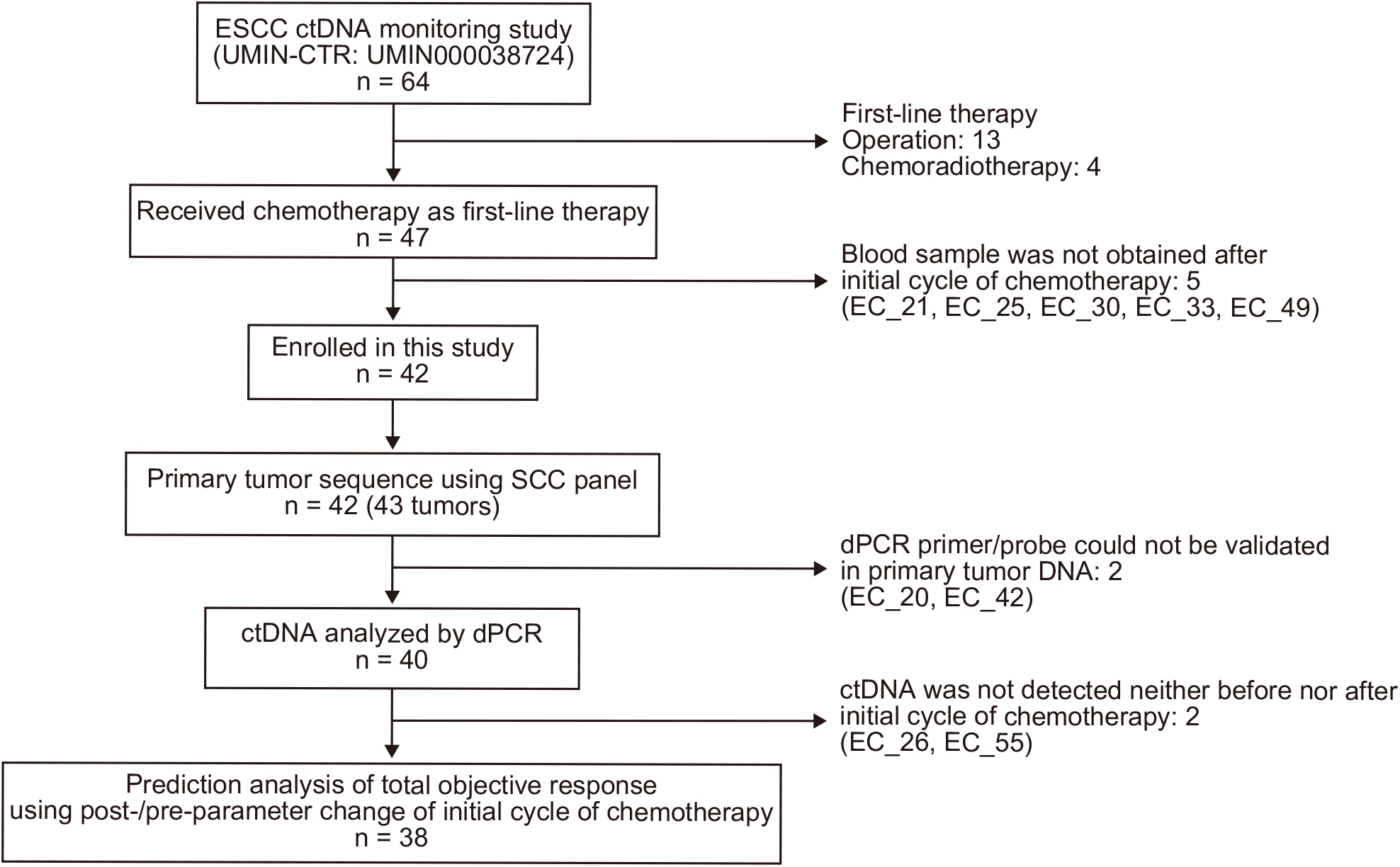
Flowchart of patients through the study and reasons for exclusion. UMIN-CTR, university hospital medical information network center clinical trials registry.

### Measurement of tumor volume

As it is difficult to accurately measure a tumor’s volume in the hollow, tubular organ using a CT scan, RECIST labels primary tumors of the gastrointestinal tracts as “non-target lesions” (7). In daily practice for ESCC patients, however, continuation of a treatment is often decided by the volume change of the primary tumor, because it is the largest lesion in most ESCC patients. In this study, changes in primary tumor volumes were included in an evaluation of treatment efficacy. A partial response is defined as a >30% decrease in the sum of the longest distance from baseline in RECIST and as a >50% decrease of the square measure according to the World Health Organization’s criteria (7,22). These are converted to a >65% decrease of the volume of a sphere. An esophageal primary tumor often extends along the longitudinal axis and forms a vertically long ellipsoid. The longest distance or square measure of a cross section of the primary tumor may not reflect the actual tumor volume. It is more reliable to approximate the volume using a surface area measurement on a CT scan, but this requires significant investments in time and software. It has been reported that the ellipsoidal approximation of volume using 3 measured axes closely estimates the measured tumor volume from a volumetric analysis completed using the available software (23). In this study, therefore, the tumor volume was measured by ellipsoidal approximation. The first 2 half-axes (*a* and *b*) were obtained by measuring the diameters on the axial scan where the lesion was the most visible. The last half-axis (*c*) was obtained by measuring the height of the lesion, calculated as by multiplying the slice thickness by the number of slices in which the tumor was visible on the axial scan. The volume of each tumor was defined as:

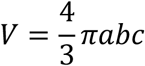

At each CT evaluation during first-line chemotherapy, all primary and metastatic tumors were measured and the volumes of all tumors were summed to calculate the “total tumor volume.” Lesions diagnosed as metastasizing at any time point during the disease course were chronologically evaluated, and lesions with a diameter over 5 mm were measured.

### Panel sequencing

Tumor- and corresponding PBMC-DNA samples were subjected to amplicon sequencing using the Ion Proton™ in 34 patients and the Ion S5™ system in 9 patients (Supplementary Methods). In the esophagus of EC_38, 2 primary lesions were observed, labeled EC_38_T1 and EC_38_T2, which were analyzed by Ion Proton™ and Ion S5™, respectively. Samples were sequenced using a specifically designed SCC panel covering 617 exons of 31 genes that are frequently altered in ESCC and head and neck squamous cell carcinomas (Supplementary Table S2). The entire length of the panel sequence is 220.57 kb, and the average amplicon size is 202 bp.

Approximately 20 ng of DNA per sample were used to prepare barcoded libraries with IonXpress barcoded adapters and an Ion AmpliSeq Library Kit plus (Thermo Fisher Scientific). Pooled barcoded libraries were subsequently conjugated with sequencing beads by emulsion PCR and enriched on an Ion Chef using the Ion PI Hi-Q Chef kit or Ion 540 Kit Chef according to manufacturer’s instructions (Thermo Fisher Scientific). Finally, sequencing was performed using Ion PI v3 chips on Ion Proton or Ion 540 chips on Ion S5 System (Thermo Fisher Scientific).

### Identification of somatic mutations

For reference, we used either the GRCh37/hg19 or GRCh38/hg38 human genome. Alignment to the reference genomes and sequencing read counting were performed in Torrent Suite version 5.0 (Thermo Fisher Scientific), Hierarchical Indexing for Spliced Alignment of Transcripts (Hisat2), and Burrows-Wheeler Aligner (BWA). Somatic mutations, including single-nucleotide variants, insertions, and deletions, were detected using a variant call algorithm in tumor- and matched-PBMC samples from the Ion Reporter software 5.0 tumor-normal workflow (Thermo Fisher Scientific), as previously described (24), or VarScan2 (25) in which germline variants were subtracted from the tumor variants. In those cases, analyzed using the 3 algorithms, commonly detected mutations were prioritized in the following mutation selection.

Using Integrative Genomics Viewer software, all identified single-nucleotide variants, insertions, and deletions were visually inspected to filter out possible strand-specific errors, such as a mutation detected in only the forward or reverse DNA strand. The dbSNP database was used to exclude single-nucleotide polymorphisms from the called variants. The following criteria were used as cutoffs: (i) total coverage > 200; (ii) variant coverage > 10; and (iii) variant frequency > 10%. Mutations were called if they occurred in <0.1% of reads in the normal control (minor allele frequency) and were absent from the dbSNP, as well as the 1000 Genomes Project database.

### Monitoring of ctDNA levels using dPCR

The dPCR assay for quantitative monitoring of ctDNA levels was performed as described previously (21). Briefly, specific primers and probes labeled for wild-type and mutant alleles were specifically designed for each mutation identified in a primary tumor, using Hypercool Primer & Probe™ technology (Nihon Gene Research Laboratories, Sendai, Japan). We prioritized 1–3 mutations per patient with a variant allele frequency (VAF) higher than 10% in primary tumors for dPCR analysis. The criteria for mutation selection for ctDNA monitoring using dPCR are described in the Supplementary Methods, and the definitions of positive and negative ctDNA were described previously (21). VAFs of ctDNA data were plotted on a time course, along with the therapy types and total tumor volumes.

### Evaluation and prediction analysis of the chemotherapy response

For ESCC patients, it seemed that a >50% reduction in tumor volume was sufficient to continue with chemotherapy in daily practice, even though this threshold does not meet the >65% volume reduction for partial responses that was specified by RECIST or the World Health Organization’s criteria. In this study, we evaluated the objective response (OR) at the end of first-line chemotherapy based on the total tumor volume from the final CT before a change of treatment. We defined a >50% reduction of the total tumor volume at the end of the first-line chemotherapy from baseline to indicate a “total positive OR,” whereas a ≤50% reduction in the total tumor volume was labeled as a “total negative OR.” We evaluated the prediction accuracy of the total OR based on the ratio of pre- and post-treatment values of the initial cycle of chemotherapy in each parameter: post-/pre-ctDNA, post-/pre-volume, and post-/pre-TMs (-SCC, -CEA, and -CYFRA).

### Statistical analysis

To evaluate the performance of each independent factor in predicting the chemotherapy response, a receiver-operating characteristic (ROC) curve analysis was performed, with optimal cutoff values obtained from the Youden’s index (26). Diagnostic accuracy was expressed as the sensitivity, specificity, and area under the curve (AUC). For group comparisons, we used the Jonckheere-Terpstra test, Wilcoxon signed-rank test, Fisher’s exact test, and Mann-Whitney U test. Correlations between 2 variables were calculated based on Spearman’s rank correlation coefficient. Kaplan-Meier estimates with log-rank tests were used to compare OS, stratified based on the post-/pre-ctDNA level. A Cox proportional hazards model was used to estimate risks, based on OS. The post-/pre-ctDNA level was divided into 2 categories using an optimal cutoff value for stratified OS group comparisons. We considered *P* values < 0.05 to be statistically significant for all analyses. All analyses were performed using GraphPad Prism 8 (GraphPad) and EZR (27). The target sample size was set at 35 (Supplementary Methods). A total of 42 patients were enrolled in this study.

## Results

### Mutations in primary ESCC

The 42 patients enrolled in this study had diagnoses of ESCC Stages II, III, and IV (n = 4, 12, and 26, respectively). DCF, CF, and irinotecan/cisplatin were administered as first-line therapies in 37 patients, 4 patients, and 1 patient, respectively. Detailed clinical information for all 42 patients are shown in **Fig. 2A** and Supplementary Table S1. Patient EC_38 had 2 primary tumors: an SCC in the middle thoracic esophagus and a neuroendocrine carcinoma in the upper-middle thoracic esophagus. Consequently, 43 tumors had mutation screening performed using the SCC panel. In the sequence analysis of the primary tumor- and PBMC-DNA, the average base coverage depth was 2,768 and the average percentage of reads on the target was 94.5%. We identified a total of 96 mutations (2.2 mutations per sample [10.0/Mb], on average) that had a VAF >10% (Supplementary Table S3). The patient characteristics and mutation profile are summarized in **Fig. 2A**. Sequence data were deposited in the DNA Data Bank Japan (accession number JGAS00000000219) (28).

**Figure 2.**
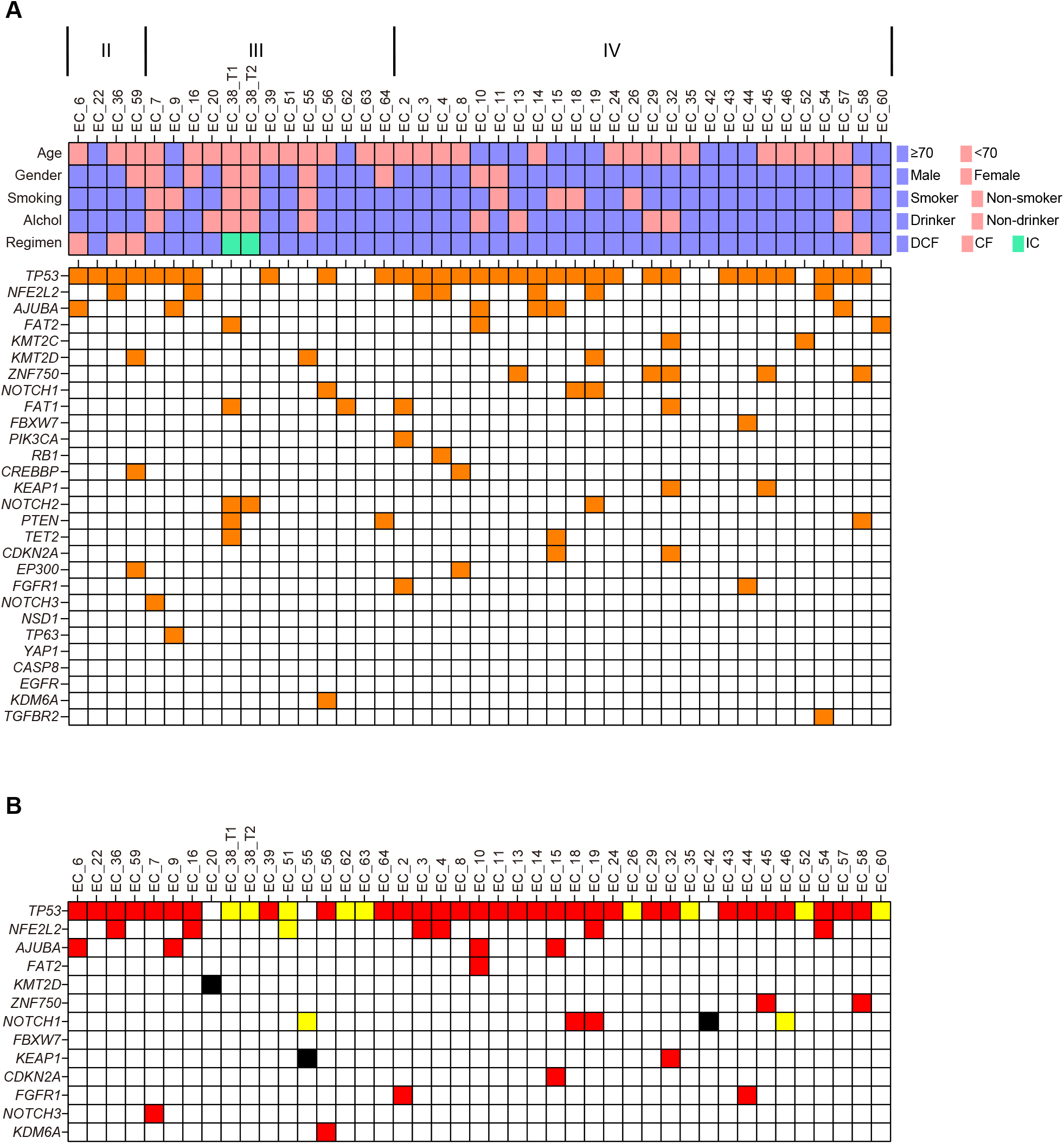
Somatic mutation profile of primary ESCC tumors form 42 patients. (A) Mutation profile of 43 ESCC tumors. Clinical characteristics are shown in the top panel. Mutated genes (>10% VAF) are shown in the bottom panel. (B) Mutations used for dPCR analysis. Red boxes indicate mutations that met the criteria for ctDNA analysis using dPCR. Yellow and black boxes represent additional mutations with specific dPCR primer/probe sets that were designed for ctDNA analysis. Yellow boxes are validated primer/probe sets using primary tumor DNA. Black boxes are primer/probe sets could not be validated using primary tumor DNA.

### CtDNA detection

In 2 cases (EC_20 and EC_42), we could not validate a probe/primer for tumor-specific mutations. Therefore, probe/primer sets for 70 selected mutations from 40 patients were validated by dPCR using primary tumor DNA (**Fig. 2B**; Supplementary Table S3). Out of the 40 patients, those with more advanced disease stages had higher VAFs in pretreatment plasma (*P* = 0.002; Jonckheere-Terpstra test; **Fig. 3A**). There were 2 patients (EC_26, Stage IVA; EC_55, Stage III) with negative ctDNA levels. Overall, 95% (38/40) of Stage II or higher patients had positive pretreatment ctDNA levels. The total tumor volume was also correlated with the pretreatment ctDNA VAF (*r* = 0.60; 95% confidence interval [CI], 0.35–0.77; *P* < 0.001; Spearman’s rank correlation coefficient; **Fig. 3B**).

**Figure 3.**
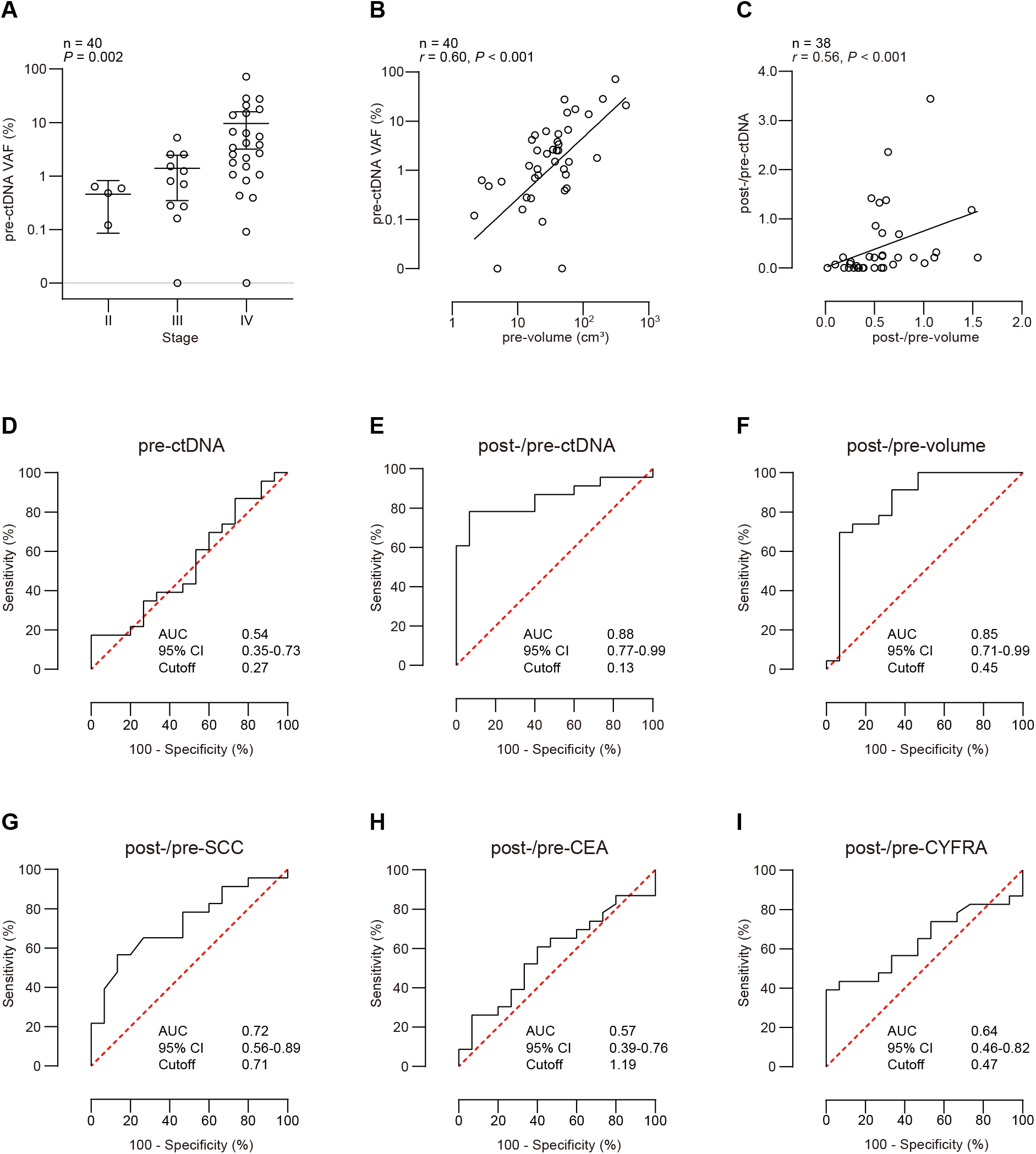
Pretreatment levels and early changes in parameters for predictions of later responses to chemotherapy. (A) VAF of ctDNA in pretreatment plasma in terms of TNM stage. (B) Correlation between VAF of ctDNA in pretreatment plasma and pretreatment total tumor volume. (C) The association between post-/pre-ctDNA and post-/pre-volume. (D–I) ROC curve analyses of predictions for later responses to chemotherapy by (D) pretreatment ctDNA, (E) post-/pre-ctDNA, (F) post-/pre-volume, post-/pre-SCC, (H) post-/pre-CEA,and (I) post/pre-CYFRA.

### Early changes in ctDNA levels, tumor volumes, and tumor markers during chemotherapy

In patients EC_26 and EC_55, ctDNA was not detected in blood samples taken before and after an initial cycle of chemotherapy (**Fig. 1**). Consequently, the early ctDNA change during chemotherapy was evaluated in 90.5% (38/42) of enrolled patients. The correlation between post-/pre-ctDNA and post-/pre-volume was not strong (*r* = 0.56; 95% CI, 0.29– 0.75; *P* < 0.001; Spearman’s rank correlation coefficient; **Fig. 3C**). This result indicates a discrepancy between early ctDNA changes and early tumor volume changes in predictions of later chemotherapy responses. Correlations between post-/pre-TMs and post-/pre-ctDNA levels were not observed (Supplementary Fig. S1).

### Prediction of chemotherapy response using early changes in ctDNA

Among the 38 patients evaluated, 23 (60.5%) showed a total positive OR at the end of first-line chemotherapy, while 15 (39.5%) showed a total negative OR. According to the ROC curve analysis, the prediction accuracy for the total OR by pretreatment ctDNA level was low in cases of advanced ESCC. The AUC was 0.54 (95% CI, 0.35–0.73). The optimal cutoff value of pretreatment ctDNA was 0.27, at which point the sensitivity was 0.17 and the specificity was 1.00 (**Fig. 3D**). Meanwhile, post-/pre-ctDNA levels predicted the total OR with high accuracy in cases of advanced ESCC. The AUC was 0.88 (95% CI, 0.77–0.99). The optimal cutoff value of post-/pre-ctDNA was 0.13, at which point the sensitivity was 0.78 and the specificity was 0.93 (**Fig. 3E**). In terms of tumor volume changes measured by CT after an initial cycle of chemotherapy, the ROC curve analysis revealed that the AUC of the post-/pre-volume for predictions of total ORs was 0.85 (95% CI, 0.71–0.99). The optimal cutoff value for the post-/pre-volume was 0.45, at which point the sensitivity was 0.74 and the specificity was 0.93 (**Fig. 3F**). The AUC values of post-/pre-SCC, post-/pre-CEA, and post-/pre-CYFRA were lower (0.72, 0.57, and 0.64, respectively) than those of post-/pre-ctDNA and post-/pre-volume (**Fig. 3G–I**). Among blood biomarkers, only the AUC of early ctDNA changes was equivalent to that of early tumor volume changes as seen on CT images.

### Dynamics of ctDNA and tumor volume during chemotherapy for ESCC patients

Results of dynamics of ctDNA levels and tumor volumes during chemotherapy are illustrated in **Fig. 4** and Supplementary Fig. S2. Patients with levels of post-/pre-ctDNA that were below the cutoff value of 0.13 were assigned to the low post-/pre-ctDNA group (n = 18), whereas those with values above the cutoff were assigned to the high post-/pre-ctDNA group (n = 20). Out of 18 patients, 17 (94.4%) with low post-/pre-ctDNA levels showed a total positive OR (**Fig. 4A** and **B**; Supplementary Fig. S2A). In the remaining patient (EC_3), although we observed significant reductions of both the tumor volume and ctDNA level after an initial cycle of DCF, cancer treatment could not be continued after the second cycle due to the adverse effect of interstitial pneumonia (Supplementary Fig. S2A). Meanwhile, 14 out of 20 (70.0 %) patients with high post-/pre-ctDNA levels showed total negative ORs (**Fig. 4C** and **D**; Supplementary Fig. S2B). The remaining 6 patients with high post-/pre-ctDNA levels showed total positive ORs (Supplementary Fig. S2B).

**Figure 4.**
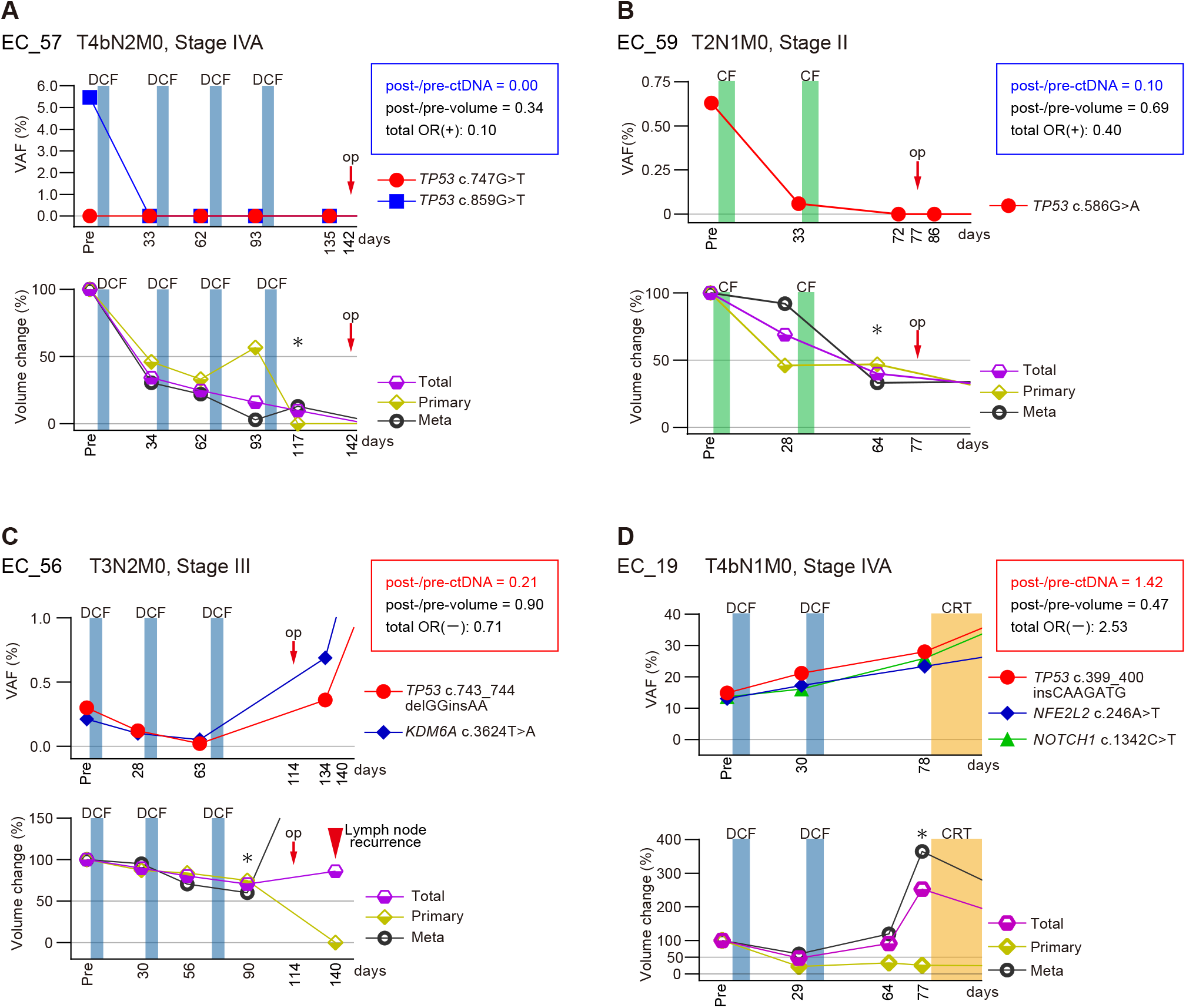
Dynamics of ctDNA and tumor volume during first-line chemotherapy of ESCC. Representative cases classified low post-/pre-ctDNA, (A) EC_57 and (B) EC_59, and high post-/pre-ctDNA, (C) EC_56 and (D) EC_19, were shown. Asterisk indicates the timepoint of the CT in which the total OR was assessed. CRT, chemoradiotherapy; op, operation.

Although the correlation between post-/pre-ctDNA levels and post-/pre-volumes was weak (**Fig. 3C**), the AUC of post-/pre-ctDNA levels in the prediction of the total OR was equivalent to that of post-/pre-volumes (**Fig. 3E** and **F**). Next, the association between post-/pre-ctDNA levels and total OR was evaluated according to the early tumor volume change after an initial cycle of chemotherapy. Out of 38 patients, 18 (47.4%) had a post-/pre-volume <50% and 20 (52.6%) had a post-/pre-volume ≥50%. We observed significant decreases of VAFs in post-ctDNA from baseline in patients with a post-/pre-volume <50%, but not in those with a post-/pre-volume ≥50% (*P* = 0.001 and *P* = 0.070, respectively; Wilcoxon signed-rank test; **Fig. 5A**). In the group with post-/pre-volumes <50%, 13 of 18 (72.2%) patients showed low post-/pre-ctDNA levels. Out of the 13 patients with low post-/pre-ctDNA levels, 12 (92.3%) showed a total positive OR. We did not observe a significant difference in the numbers of total positive and negative OR patients according to the post-/pre-ctDNA level (*P* = 0.11; Fisher’s exact test; **Fig. 5B**). Among 5 patients with high post-/pre-ctDNA levels despite post-/pre-volumes <50%, 1 patient (EC_19) showed rapid disease progression despite a temporal tumor volume reduction after an initial cycle of DCF. In this case, the tumor growth might have been predicted by an early ctDNA change, as continuous elevation of the ctDNA level was observed throughout the disease course (**Fig. 4D**).

**Figure 5.**
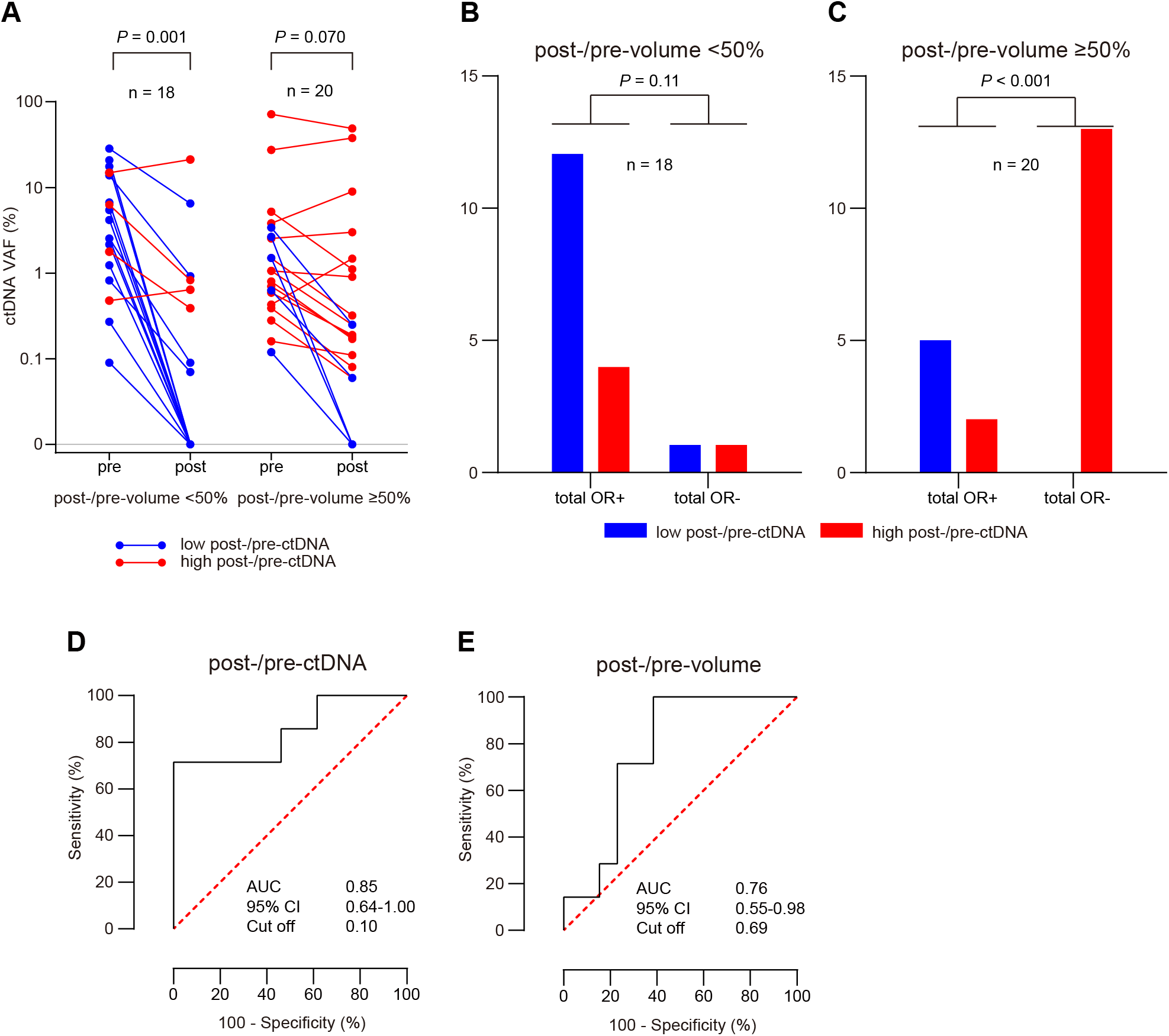
Later response of chemotherapy by early change of ctDNA and tumor volume. Change of ctDNA VAF before and after initial cycle of chemotherapy in patients with post-/pre-volume <50% and ≥ 50%. (B and C) Number of patients showed total positive and negative ORs according to post-/pre-ctDNA status in patients with post-/pre-volume (B) <50% and (C) ≥50%. (D and E) ROC curve analysis of prediction for later response for chemotherapy in patients with post-/pre-volume ≥50% by (D) post-/pre-ctDNA and (E) post-/pre-volume.

In contrast, in the group with post-/pre-volumes ≥50%, 13 out of 15 (86.7%) patients with high post-/pre-ctDNA levels showed total negative ORs (**Fig. 5C**). However, all 5 of the patients with low post-/pre-ctDNA levels showed total positive ORs, although tumor shrinkage was not observed after an initial cycle of chemotherapy (**Fig. 5C**). These results indicate that among the patients evaluated as having SD by CT after an initial cycle of chemotherapy, most patients with high post-/pre-ctDNA levels did not achieve remarkable tumor shrinkage by continuation of chemotherapy. Meanwhile, patients with low post-/pre-ctDNA levels had good responses to chemotherapy even if their post-/pre-volumes were ≥50%. A significant difference was observed in the numbers of total positive and negative OR patients according to the post-/pre-ctDNA levels in the group with post-/pre-volumes ≥50% (*P* < 0.001; Fisher’s exact test; **Fig. 5C**). Of note, in patients with post-/pre-volumes ≥50%, the AUC of post-/pre-ctDNA levels for predictions of total ORs was higher than that of post-/pre-volumes (0.85 [95% CI, 0.64–1.00] vs 0.76 [95% CI, 0.55–0.98], respectively; **Fig. 5D** and **E**).

### Duration of response for chemotherapy and overall survival by early ctDNA change

**Fig. 6A** and **B** plot tumor volume changes until 20 months in patients with post-/pre-volumes <50% and ≥50%, respectively. In both groups, patients with low post-/pre-ctDNA levels generally had tumor volumes below 50% throughout the disease course. Details of the study patients treatments and outcomes are shown in Supplementary Table S4. Positive and negative ctDNA levels and clinical information for patients are schematized in Supplementary Fig. S3. We also evaluated the association between early ctDNA changes and the duration of response (DOR) to chemotherapy. In this study, the DOR was defined as the length of time until confirmation of a >50% total tumor volume reduction during the first-line chemotherapy. Patients with low post-/pre-ctDNA levels (n = 18) had longer DORs than patients with high post-/pre-ctDNA levels (n = 20; 71.7 ± 48.0 days vs 29.8 ± 45.4 days, respectively; *P* = 0.001; Mann-Whitney U test; **Fig. 6C**). By comparing the pretreatment ctDNA status with the ctDNA decrease within 3 months of the start of first-line treatment, our previous study of ctDNA monitoring for ESCC patients demonstrated that treatment efficacy was a greater factor for OS than the pretreatment tumor burden (21). The present study also demonstrated that patients with low post-/pre-ctDNA levels (n = 18) showed significantly better OS than those with high post-/pre-ctDNA levels (n = 20; hazard ratio, 0.40; 95% CI, 0.17–0.93; *P* = 0.031; log-rank test; **Fig. 6D**). These results indicate that early ctDNA dynamics after an initial cycle of chemotherapy may predict not only later responses and DORs, but also OS.

**Figure 6.**
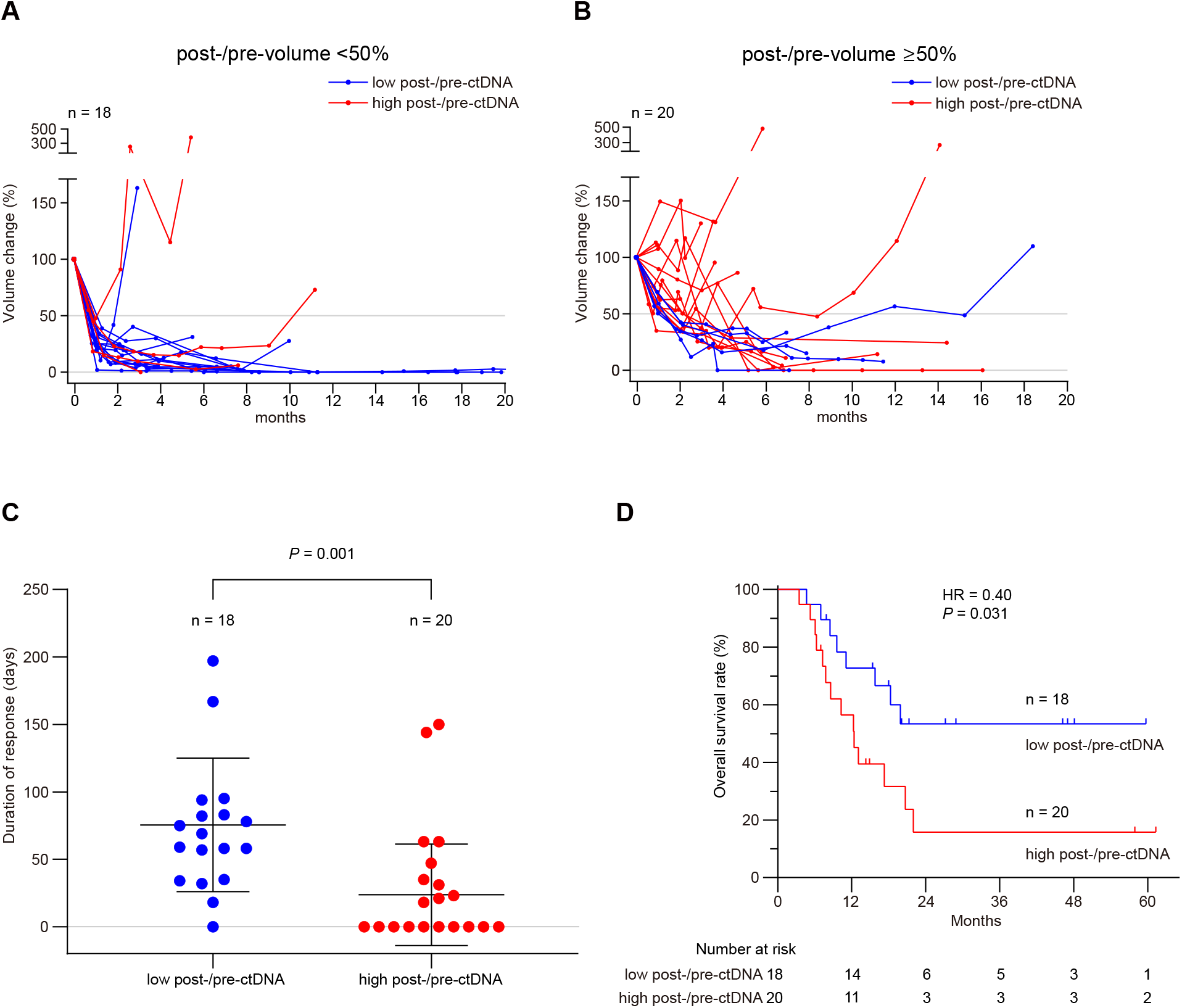
Medium- to long-term outcomes of treatments for ESCC patients according to post-/pre-ctDNA status. (A and B) Dynamics of total tumor volume until 20 months from start of treatment in patients with post-/pre-volume (A) <50% and (B) ≥50%. (C) Duration of response for first-line chemotherapy. (D) Overall survival stratified by post-/pre-ctDNA status.

## Discussion

In this study, we have demonstrated that early ctDNA changes before and after an initial cycle of chemotherapy could predict later responses after several cycles in ESCC patients. The optimal time point for the evaluation of chemotherapy responses using early ctDNA dynamics has been investigated in several studies (10,14,16). In biweekly oxaliplatin- or irinotecan-based chemotherapy for colorectal cancer, no significant difference was observed between the pretreatment ctDNA level and the level 3 days after the start of chemotherapy. However, a significant reduction in the ctDNA level before a second cycle (i.e., 2–3 weeks after treatment start) was correlated with a radiographic response at 8–10 weeks (10). In a study of 3-week cycles of capivasertib or placebo in combination with paclitaxel therapy for estrogen receptor-positive breast cancer, the start of the second cycle (i.e., 4-weeks after treatment start) was an optimal time point for measuring the ctDNA level for comparison with the day-8 level in order to predict progression-free survival (16). These reports indicated that ctDNA dynamics should be evaluated over an interval of several weeks from the start of chemotherapy. In advanced ESCC, high-dose cisplatin-based chemotherapies are usually administered early as standard therapies. Because these regimes take 3–4 weeks per cycle and early progression is often observed in ESCC, a CT evaluation is usually performed in each cycle. In our present study, post-ctDNA levels evaluated before the start of the second cycle corresponded to those measured at about 3–5 weeks after the start of the initial cycle of chemotherapy. In ESCC chemotherapy, this time point resulted in a high AUC for predictions of chemotherapy responses using post-/pre-ctDNA levels.

In previous reports on early ctDNA dynamics during chemotherapy in other cancer types, tumor responses were evaluated 8–12 weeks after the start of treatment by image diagnoses according to RECIST (10,13,16,17). To supplement these reports in other malignancies, this study examined the difference between early dynamics of tumor volume by ellipsoidal approximation and ctDNA after an initial cycle of chemotherapy. For predicting the total OR, the AUC of the post-/pre-ctDNA level was equivalent to that of post-/pre-volumes (0.88 vs 0.85, respectively; **Fig. 3E** and **F**). The evaluation of total tumor volume by ellipsoidal approximation is a bit complicated, as is the volumetric analysis, because (i) many cases have multiple metastatic lesions; (ii) distinguishing between metastasized lymph nodes and nonspecific node swelling is often difficult; (iii) many cases have indistinct borders between tumor and normal tissue, especially in primary tumors; and (iv) distinguishing between remnants of a tumor and scar tissue after treatment is often difficult. The calculation of ctDNA changes to reflect changes in all lesions is convenient and objective. Early ctDNA changes will be a great help in evaluating and predicting chemotherapy efficacy in daily practice for ESCC.

Chemotherapy is usually continued until progressive disease is evident by image diagnosis in ESCC patients. It seems that prediction of chemotherapy efficacy by post-/pre-ctDNA levels may be useful for patients diagnosed as having SD without significant tumor reduction or progression after an initial cycle. Among 20 patients with post-/pre-volumes ≥50%, 15 had high post-/pre-ctDNA levels, of which 13 (86.7%) showed total negative ORs after chemotherapy (**Fig. 5C**). In contrast, all 5 patients with low post-/pre-ctDNA levels showed total positive ORs, although sufficient tumor volume reduction was not observed after an initial cycle. Consequently, the total OR could be predicted by the post-/pre-ctDNA data in 18 out of 20 (90%) patients. The AUC for the total OR prediction by post-/pre-ctDNA levels was higher than that by post-/pre-volumes in patients with post-/pre-volumes ≥50% (0.85 vs 0.76, respectively; **Fig. 5D** and **E**), although this difference was not observed when evaluating all 38 patients (**Fig. 3E** and **F**). These results indicate that post-/pre-ctDNA levels could predict responses at the end of the chemotherapy even in patients who have been evaluated as having SD by CT after an initial cycle of chemotherapy.

It has been reported that a reduction in the ctDNA level of more than 80–90% after 1 cycle is correlated with the tumor response, based on RECIST criteria, in colorectal cancer patients (10,13). In the present study, the optimal cutoff value for the post-/pre-ctDNA level in predicting the total OR was 0.13 (87% reduction; **Fig. 3E**). Kurtz et al. (14) investigated early ctDNA dynamics during chemotherapy and assessed the end-of-therapy response by PET/CT in diffuse large B-cell lymphoma. The study demonstrated that patients achieving a complete response had a large drop in ctDNA, with a 2-log decrease after 1 cycle (14). In the present study, all 13 patients with a 2-log decrease after the initial cycle (i.e., post-/pre-ctDNA, 0) also showed a total positive OR (**Fig. 5A–C**). These results indicate that a >90% reduction of ctDNA after an initial cycle may result in a good response if the chemotherapy is continued, regardless of the cancer type.

Our study also demonstrated that patients with low post-/pre-ctDNA levels had longer DORs and showed better OS than those with high post-/pre-ctDNA levels (**Fig. 6A–D**). In this study, patients continued chemotherapy according to the conventional evaluation by CT as a daily practice for ESCC. It was suggested that an early assessment of chemotherapy efficacy by ctDNA change would enable an earlier switch to an alternate therapy, allowing a patient with no response to chemotherapy to receive more effective treatment earlier. A recent meta-analysis demonstrated that even a 4-week delay of cancer treatment was associated with increased mortality in several major cancer types (29). It is possible that earlier changes to an alternate treatment form ineffective therapy may increase the odds of survival in ESCC patients.

Considering subsequent disease progression after chemotherapy in patients with high post-/pre-ctDNA levels and the ambiguity of evaluation by CT, few patients may suffer from early treatment changes made based on the efficacy evaluation of early ctDNA changes. Nevertheless, in some patients, it may be difficult to change treatment plans using only the results of high post-/pre-ctDNA levels, despite detecting tumor shrinkage. However, a ctDNA test can be easily repeated over short intervals, such as a week, because our ctDNA monitoring system using dPCR offers a rapid turnaround time of as little as 1 day and has an affordable cost (21). Sustained high ctDNA levels in retests may indicate a lack of response to chemotherapy more clearly than using only post-/pre-ctDNA levels from 2 time points, before and after the initial cycle.

The small sample size of our present study impairs the generalization of our conclusions. Randomized controlled trials with larger cohort of patients are needed to prove the clinical utility of early ctDNA changes for predictions of chemotherapy efficacy. However, information on early ctDNA dynamics can add an objective index of disease control or progression during chemotherapy to supplement conventional image diagnoses.

In conclusion, early ctDNA changes before and after an initial cycle of chemotherapy predict later responses at the end of chemotherapy with high accuracy in ESCC patients.

## Data Availability

Sequence data were deposited in the DNA Data Bank Japan (Accession number JGAS00000000219).

