## Supplementary Files for "Early dynamics of circulating tumor DNA predict chemotherapy responses for patients with esophageal cancer"

#### Supplementary Information

**Supplementary Methods**
**Supplementary Table S1.** Clinicopathological characteristics of the study patients
**Supplementary Table S2.** Target genes in the specifically designed SCC panel
**Supplementary Table S3.** Primary tumor mutations detected by sequence analysis and circulating tumor DNA levels measured by digital PCR
**Supplementary Table S4.** Treatments and outcomes of the study patients
**Supplementary Figure S1.** Correlation between early change of ctDNA and conventional tumor markers.
**Supplementary Figure S2.** Dynamics of ctDNA and tumor volume during first-line chemotherapy in the study patients.
**Supplementary Figure S3.** Longitudinal ctDNA monitoring and clinical information.

This supplementary material provides additional information about this study that was not included in the main text.

### Supplementary Methods

#### *Identification of somatic mutations in primary tumor tissue*

Among 43 tumors from 42 patients, the first 34 tumors and their corresponding peripheral blood mononuclear cell (PBMC) DNA were analyzed by an Ion Proton$^{TM}$ system as previously described (reference 21 in the main text). The later 9 tumors and corresponding PBMC DNA were analyzed using an Ion S5$^{TM}$ system. The sequence platforms used in each case are listed in Supplementary Table S3. In the later 9 tumors, alignment to the reference genomes and sequencing read counting were performed in Torrent Suite version 5.0 (Thermo Fisher Scientific), Hisat2 (https://ccb.jhu.edu/software/hisat2/index.shtml), and BWA（Burrows–Wheeler Alignment tool). In those cases, analyzed using the 3 algorithms, commonly detected mutations were prioritized in the following mutation selection, as described in the main text. In terms of insertion and deletion mutations, the following criteria were used as cutoffs: (i) total coverage > 200; (ii) variant coverage > 10; and (iii) variant frequency > 20%.

#### *The criteria for mutation selection in ctDNA monitoring using digital PCR*

According to our previous study, between 1 and 3 mutations, with variant allele frequencies (VAFs) >10%, found in primary tumors were prioritized for digital PCR (dPCR) analysis (reference 21 in the main text). The criteria used to select mutations for circulating tumor DNA (ctDNA) monitoring using dPCR were: (a) the mutation having the highest VAF in the primary tumor; (b) up to additional 2 mutations with a VAF >10% in primary tumors; (c) recurrent mutations observed among patients; (d) mutated genes having a high success rate for primer/probe validation; and (e) exclusion of mutations with a surrounding sequence suspected of having low PCR efficiency and low specificity of hybridized probes (e.g., self-complementarity and/or secondary structure of the primer/probe). Furthermore, those mutations already validated in tumor DNA from ESCC or other types of malignancies could reduce the temporary cost and time needed for syntheses of specifically designed probes for each tumor-specific mutation. Therefore, mutations listed in our dPCR primer/probe library were analyzed, even if the mutations detected in primary tumors were not matched to the criteria for mutation selection.

#### *Longitudinal ctDNA monitoring and clinical information during the treatment course*

Longitudinal binarized (i.e., ctDNA-positive and -negative) ctDNA status and clinical information from during and after first-line chemotherapy in 17 patients, among the 38 with ctDNA evaluated, are schematized in Supplementary Figure S3. The information from the remaining 21 patients has been previously published (reference 21 in the main text).

##### *Sample size estimation*

In a survival analysis from our previous study, the 1-year overall survival (OS) rate for patients with a ctDNA decrease within 90 days was 90% (n = 11), while the OS rate of patients with a sustained positive ctDNA level was 27% (n = 10; reference 21 in the main text). The proportion of patients with a total positive OR rate, with or without a ctDNA decrease after an initial chemotherapy cycle, was expected to be almost the same as the proportion that survived to 1 year. With a 60% difference between 2 groups, the target sample size needed to be 26 patients (alpha-error, 0.05; power, 0.8) for both groups combined. In an ROC analysis, the sample size was required to be 28 (alpha-error, 0.05; power, 0.9, and area under the curve, 0.8). In our previous study, ctDNA could not be evaluated in 5% of patients, due to a lack of adequate tumor samples for NGS analyses or specific dPCR probe/primers validated in tumor DNA. Furthermore, the pretreatment rate of negative ctDNA levels in ESCC patients with Stage II or higher was 15%. Therefore, the target sample size estimate was 35.

**Supplementary Table S1. Clinicopathological characteristics of the study patients**

| Patient ID | Age range | Gender | Locus [a] | UICC Stage [b] | TNM [b] | Pretreatment total tumor volume (cm³) |
|---|---|---|---|---|---|---|
| EC_2 | 66-70 | Male | LtAeG | IVA | T4bN1M0 | 448.9 |
| EC_3 | 66-70 | Male | LtAe | IVB | T3N2M1 | 121.2 |
| EC_4 | 61-65 | Male | CeUt | IVA | T4bN1M0 | 20.1 |
| EC_6 | 56-60 | Male | Ut | II | T3N0M0 | 5.7 |
| EC_7 | 61-65 | Female | MtUt | III | T3N1M0 | 20.7 |
| EC_8 | 66-70 | Male | MtUtLt | IVA | T4bN2M0 | 50.7 |
| EC_9 | 81-85 | Male | MtLtAe | III | T3N2M0 | 19.9 |
| EC_10 | 71-75 | Female | Mt | IVA | T4bN1M0 | 28.0 |
| EC_11 | 71-75 | Female | MtUtLt | IVB | T3N3M1 | 311.2 |
| EC_13 | 81-85 | Male | Lt | IVB | T3N1M1 | 27.1 |
| EC_14 | 61-65 | Male | UtMt | IVA | T4bN2M0 | 56.0 |
| EC_15 | 76-80 | Male | MtLt | IVB | T4bN2M1 | 60.4 |
| EC_16 | 51-55 | Female | Lt | III | T3N2M0 | 18.4 |
| EC_18 | 76-80 | Male | Lt | IVB | T1bN2M1 | 16.5 |
| EC_19 | 71-75 | Male | LtAe | IVA | T4bN1M0 | 57.2 |
| EC_20 | 56-60 | Male | Ae | III | T3N1M0 | 68.7 |
| EC_22 | 71-75 | Male | Mt | II | T2N1M0 | 2.2 |
| EC_24 | 51-55 | Male | UtMt | IVA | T4bN0M0 | 34.7 |
| EC_26 | 61-65 | Male | Mt | IVA | T4bN1M0 | 4.7 |
| EC_29 | 61-65 | Male | Mt | IVA | T4bN2M0 | 40.6 |
| EC_32 | 66-70 | Male | LtMt | IVA | T4aN1M0 | 39.5 |
| EC_35 | 61-65 | Male | Mt | IVA | T4bN3M0 | 52.3 |
| EC_36 | 61-65 | Male | Ut | II | T2N0M0 | 3.6 |
| EC_38_T1 | 61-65 | Female | Mt | III | T3N2M0 | 37.2 |
| EC_38_T2 | | | MtUt | III | T3N2M0 | |
| EC_39 | 61-65 | Male | MtLt | III | T3N2M0 | 15.0 |
| EC_42 | 71-75 | Male | MtLtAe | IVB | T3N3M1 | 67.8 |
| EC_43 | 76-80 | Male | UtMt | IVA | T4bN2M0 | 23.9 |
| EC_44 | 71-75 | Male | MtUt | IVA | T4bN2M0 | 52.0 |
| EC_45 | 66-70 | Male | UtMt | IVB | T4bN3M1 | 76.2 |
| EC_46 | 56-60 | Male | MtLt | IVA | T4bN1M0 | 163.3 |
| EC_51 | 66-70 | Male | Ce | III | T3N1M0 | 16.0 |
| EC_52 | 56-60 | Male | MtLt | IVA | T4bN3M0 | 42.4 |
| EC_54 | 61-65 | Male | LtAe | IVA | T3N3M0 | 54.2 |
| EC_55 | 56-60 | Female | MtLtAe | III | T3N2M0 | 45.3 |
| EC_56 | 46-50 | Male | Lt | III | T3N2M0 | 13.7 |
| EC_57 | 46-50 | Male | Ut | IVA | T4bN2M0 | 41.7 |
| EC_58 | 71-75 | Female | Mt | IVB | T1bN3M1 | 200.4 |
| EC_59 | 56-60 | Female | Mt | II | T2N1M0 | 2.8 |
| EC_60 | 71-75 | Male | MtLt | IVA | T4bN2M0 | 58.9 |
| EC_62 | 71-75 | Male | Ut | III | T3N2M0 | 18.4 |
| EC_63 | 66-70 | Male | MtLt | III | T3N1M0 | 41.8 |
| EC_64 | 46-50 | Female | Lt | III | T3N2M0 | 11.9 |

Ae, abdominal esophagus;Ce, cervical esophagus; Lt, lower thoracic esophagus; Mt, middle thoracic esophagus; Ut, upper thoracic esophagus.

[a] Anatomical regions of the esophagus (Japanese Classification of Esophageal Cancer, 11th edition).

[b] TNM classification of malignant tumours, 8th edition. Union for International Cancer Control.

**Supplementary Table S2. Target genes in the specifically designed SCC panel**

| | | | |
|---|---|---|---|
| *AJUBA* | *CASP8* | *CCND1* | *CDKN2A* |
| *CREBBP* | *EGFR* | *EP300* | *FAT1* |
| *FAT2* | *FBXW7* | *FGFR1* | *HRAS* |
| *KDM6A* | *KEAP1* | *KMT2C* | *KMT2D* |
| *NFE2L2* | *NOTCH1* | *NOTCH2* | *NOTCH3* |
| *NSD1* | *PIK3CA* | *PTEN* | *RB1* |
| *SOX2* | *TET2* | *TGFBR2* | *TP53* |
| *TP63* | *YAP1* | *ZNF750* | |

**Supplementary Table S3. Primary tumor mutations detected by sequence analysis and circulating tumor DNA levels measured by digital PCR**

| Case ID | Chromosomal location | Gene | Variant type | Base change | Amino acid change | Total allele | Normal allele | Variant allele | Primary VAF (%) | Sequence platform | dPCR[a] | Pre-ctDNA VAF[d] | Post-ctDNA VAF[d] |
|---|---|---|---|---|---|---|---|---|---|---|---|---|---|
| EC_2 | chr4:187549359 | *FAT1* | missense | c.4759C>G | p.Leu1587Val | 2000 | 1232 | 768 | 38.40 | Ion Proton | | | |
| | chr8:38279361 | *FGFR1* | missense | c.1128C>G | p.Asn376Lys | 1995 | 1082 | 913 | 45.76 | | yes | 19.35 | 0.00 |
| | chr17:7578235 | *TP53* | missense | c.614A>G | p.Tyr205Cys | 1999 | 1242 | 757 | 37.87 | | yes | 12.75 | 0.00 |
| | chr17:7578271 | *TP53* | missense | c.578A>G | p.His193Arg | 2000 | 1713 | 287 | 14.35 | | yes | 20.94 | 0.00 |
| | chr3:178927381 | *PIK3CA* | splicesite | c.1146-2A>G | | 1999 | 1,643 | 356 | 17.81 | | | | |
| EC_3 | chr2:178098799 | *NFE2L2* | missense | c.246A>T | p.Glu82Asp | 1131 | 665 | 466 | 41.20 | Ion Proton | yes | 13.27 | 0.89 |
| | chr17:7578431 | *TP53* | nonsense | c.499C>T | p.Gln167Ter | 277 | 118 | 159 | 57.40 | | yes | 13.80 | 0.92 |
| EC_4 | chr2:178098799 | *NFE2L2* | missense | c.246A>T | p.Glu82Asp | 1322 | 875 | 447 | 33.81 | Ion Proton | yes | 0.66 | 0.00 |
| | chr13:48916851 | *RB1* | splicesite | c.381+1G>T | | 1546 | 857 | 689 | 44.57 | | | | |
| | chr17:7578190 | *TP53* | missense | c.659A>G | p.Tyr220Cys | 1996 | 633 | 1363 | 68.29 | | yes | 1.06 | 0.25 |
| EC_6 | chr14:23451144 | *AJUBA* | frameshift insertion | c.331_332insGC | p.Leu111fs | 1995 | 1316 | 679 | 34.04 | Ion Proton | yes | 0.07 | 0.00 |
| | chr17:7578190 | *TP53* | missense | c.659A>G | p.Tyr220Cys | 1997 | 1115 | 882 | 44.17 | | yes | 0.59 | 0.19 |
| EC_7 | chr17:7577500 | *TP53* | frameshift deletion | c.780_780delC | p.Ser260fs | 1990 | 1407 | 583 | 29.30 | Ion Proton | yes | 0.80 | 0.17 |
| | chr19:15292506 | *NOTCH3* | nonsense | c.2673C>A | p.Cys891Ter | 845 | 625 | 220 | 26.04 | | yes | 0.31 | 0.20 |
| EC_8 | chr16:3842089 | *CREBBP* | missense | c.1223A>G | p.His408Arg | 1137 | 974 | 163 | 14.34 | Ion Proton | | | |
| | chr17:7578235 | *TP53* | missense | c.614A>G | p.Tyr205Cys | 1999 | 1473 | 526 | 26.31 | | yes | 1.06 | 0.91 |
| | chr22:41565575 | *EP300* | missense | c.4241A>G | p.Tyr1414Cys | 1232 | 928 | 304 | 24.68 | | | | |
| EC_9 | chr3:189604293 | *TP63* | missense | c.1460G>A | p.Arg487His | 1983 | 1726 | 257 | 12.96 | Ion Proton | | | |
| | chr14:23451000 | *AJUBA* | frameshift insertion | c.475_476insA | p.Ser159fs | 1989 | 1424 | 565 | 28.41 | | yes | 2.55 | 3.01 |
| | chr17:7578455 | *TP53* | missense | c.475G>C | p.Ala159Pro | 202 | 159 | 43 | 21.29 | | yes | 0.88 | 1.25 |
| EC_10 | chr5:150907682 | *FAT2* | missense | c.10039G>A | p.Asp3347Asn | 1991 | 1610 | 381 | 19.14 | Ion Proton | yes | 0.12 | 0.00 |
| | chr14:23450825 | *AJUBA* | frameshift insertion | c.650_651insCA | p.Gln217fs | 1991 | 1204 | 787 | 39.53 | | yes | 2.17 | 0.00 |
| | chr17:7578370 | *TP53* | splicesite | c.559+1C>T | | 1999 | 1378 | 621 | 31.07 | | yes | 1.71 | 0.00 |
| EC_11 | chr17:7577556 | *TP53* | missense | c.725G>T | p.Cys242Phe | 1988 | 1343 | 645 | 32.44 | Ion Proton | yes | 71.56 | 49.11 |
| EC_13 | chr17:7579358 | *TP53* | missense | c.329G>T | p.Arg110Leu | 1766 | 1447 | 319 | 18.06 | Ion Proton | yes | 6.28 | 0.83 |
| | chr17:80788479 | *ZNF750* | nonsense | c.1711C>T | p.Arg571Ter | 1673 | 1357 | 316 | 18.89 | | | | |
| EC_14 | chr2:178095691 | *NFE2L2* | missense | c.1640A>G | p.Asp547Gly | 1999 | 1341 | 658 | 32.92 | Ion Proton | | | |
| | chr2:178098809 | *NFE2L2* | missense | c.236A>C | p.Glu79Ala | 1995 | 1352 | 643 | 32.23 | | | | |
| | chr14:23447575 | *AJUBA* | frameshift insertion | c.1085_1086insTGGACAGCCTCTACCACACCCA | p.Gln362fs | 1975 | 795 | 1180 | 59.75 | | | | |
| | chr17:7578550 | *TP53* | missense | c.380C>T | p.Ser127Phe | 209 | 68 | 141 | 67.46 | | yes | 0.43 | 1.48 |
| EC_15 | chr17:7577120 | *TP53* | missense | c.818G>A | p.Arg273His | 1170 | 461 | 708 | 60.51 | Ion Proton | yes | 1.42 | 0.34 |
| | chr14:23450675 | *AJUBA* | frameshift insertion | c.800_801insTA | p.Gly268fs | 1766 | 697 | 974 | 55.15 | | yes | 0.69 | 0.00 |
| | chr9:21971035 | *CDKN2A* | frameshift deletion | c.310_322delCTGGACGTGCGCG | p.Leu104fs | 1857 | 1040 | 817 | 44.00 | | yes | 1.49 | 0.32 |
| | chr4:106157845 | *TET2* | nonsense | c.2746C>T | p.Gln916Ter | 1241 | 1116 | 125 | 10.07 | | | | |
| EC_16 | chr17:7578271 | *TP53* | missense | c.578A>T | p.His193Leu | 1998 | 551 | 1447 | 72.42 | Ion Proton | yes | 4.59 | 0.64 |
| | chr2:178098806 | *NFE2L2* | missense | c.239C>A | p.Thr80Lys | 1735 | 789 | 946 | 54.52 | | yes | 5.22 | 1.11 |
| EC_18 | chr17:7578235 | *TP53* | missense | c.614A>G | p.Tyr205Cys | 2000 | 1719 | 281 | 14.05 | Ion Proton | yes | 4.16 | 0.00 |
| | chr9:139391555 | *NOTCH1* | missense | c.6636C>A | p.Asp2212Glu | 1999 | 1408 | 591 | 29.56 | | yes | 1.98 | 0.00 |
| EC_19 | chr1:120539833 | *NOTCH2* | missense | c.538G>A | p.Glu180Lys | 1999 | 1870 | 362 | 18.11 | Ion Proton | | | |
| | chr2:178098799 | *NFE2L2* | missense | c.246A>T | p.Glu82Asp | 1990 | 1521 | 469 | 23.57 | | yes | 13.08 | 17.25 |
| | chr9:139412303 | *NOTCH1* | nonsense | c.1342C>T | p.Arg448Ter | 1989 | 1400 | 589 | 29.61 | | yes | 13.65 | 16.16 |
| | chr17:7578526 | *TP53* | frameshift insertion | c.399_400insCAAGATG | p.Phe134fs | 207 | 127 | 80 | 38.65 | | yes | 14.90 | 21.21 |
| | chr12:49445046 | *KMT2D* | missense | c.2420C>G | p.Ser807Cys | 391 | 173 | 74 | 18.93 | | | | |
| EC_20 | chr12:49445055 | *KMT2D* | missense | c.2411T>C | p.Leu804Ser | 1136 | 1073 | 63 | 5.55 | Ion Proton | NV[c] | | |

| | | | | | | | | | | | | | |
|---|---|---|---|---|---|---|---|---|---|---|---|---|---|
| EC_22 | chr17:7578403 | *TP53* | missense | c.527G>A | p.Cys176Tyr | 1997 | 1418 | 579 | 28.99 | Ion Proton | yes | 0.16 | 0.00 |
| EC_24 | chr17:7579389 | *TP53* | nonsense | c.298C>T | p.Gln100Ter | 1119 | 507 | 612 | 54.69 | Ion Proton | yes | 2.66 | 0.00 |
| EC_26 | chr17:7577094 | *TP53* | missense | c.844C>T | p.Arg282Trp | 283 | 277 | 6 | 2.12 | Ion Proton | yes[b] | 0.00 | 0.00 |
| EC_29 | chr17:7577104 | *TP53* | missense | c.832C>T | p.Pro278Ser | 1447 | 1203 | 241 | 16.66 | Ion Proton | yes | 3.81 | 8.98 |
| | chr17:80789818 | *ZNF750* | frameshift deletion | c.512delA | p.Lys171fs | 1989 | 1538 | 451 | 22.67 | | | | |
| | chr4:187630547 | *FAT1* | missense | c.435G>C | p.Leu145Phe | 1430 | 1023 | 407 | 28.46 | | | | |
| | chr7:151842343 | *KMT2C* | missense | c.14069G>A | p.Arg4690Gln | 1068 | 835 | 233 | 21.82 | | | | |
| | chr9:21971036 | *CDKN2A* | missense | c.322G>C | p.Asp108His | 866 | 537 | 329 | 37.99 | | | | |
| EC_32 | chr17:7577518 | *TP53* | missense | c.760_763delATCAinsTTCT | p.Ile254_Ile255delinsPhePhe | 1984 | 1532 | 446 | 22.48 | Ion Proton | yes | 0.16 | 0.00 |
| | chr17:7579315 | *TP53* | frameshift insertion | c.371_372insTG | p.Thr125fs | 1971 | 1512 | 459 | 23.29 | | yes | 2.32 | 0.00 |
| | chr17:80788986 | *ZNF750* | nonsense | c.1345G>T | p.Glu449Ter | 1355 | 771 | 584 | 43.10 | | | | |
| | chr19:10602845 | *KEAP1* | missense | c.733G>T | p.Val245Phe | 1999 | 1389 | 610 | 30.52 | | yes | 2.54 | 0.09 |
| EC_35 | chr17:7577528 | *TP53* | missense | c.743G>A | p.Arg248Gln | 1992 | 1831 | 112 | 5.60 | Ion Proton | yes[b] | 0.39 | 0.08 |
| | chr2:178098959 | *NFE2L2* | missense | c.86A>G | p.Asp29Gly | 2000 | 1477 | 523 | 26.15 | | yes | 0.00 | 0.00 |
| EC_36 | chr17:7578190 | *TP53* | missense | c.659A>G | p.Tyr220Cys | 1560 | 846 | 714 | 45.77 | Ion Proton | yes | 0.48 | 0.64 |
| | chr17:7578371 | *TP53* | missense | c.559 G>C | p.Gly187Arg | 1936 | 1503 | 405 | 20.92 | | yes | 0.00 | 0.00 |
| | chr1:120471712 | *NOTCH2* | missense | c.3779G>A | p.Arg1260His | 1989 | 1269 | 720 | 36.20 | | | | |
| | chr4:106155185 | *TET2* | missense | c.86C>G | p.Pro29Arg | 1851 | 1454 | 397 | 21.45 | | | | |
| | chr4:106196951 | *TET2* | missense | c.5284A>G | p.Ile1762Val | 1414 | 802 | 612 | 43.28 | | | | |
| EC_38-T1 | chr4:187629538 | *FAT1* | missense | c.1444G>A | p.Val482Ile | 443 | 240 | 203 | 45.82 | Ion Proton | | | |
| | chr4:187629770 | *FAT1* | missense | c.1212T>G | p.Ser404Arg | 1560 | 904 | 656 | 42.05 | | | | |
| | chr5:150945518 | *FAT2* | missense | c.2975G>A | p.Arg992Gln | 1999 | 1170 | 829 | 41.47 | | | | |
| | chr10:89692892 | *PTEN* | missense | c.376G>C | p.Ala126Pro | 1674 | 1322 | 352 | 21.03 | | | | |
| | chr17:7579416 | *TP53* | frameshift deletion | c.250delG | p.Ala84fs | 1861 | 1587 | 274 | 14.72 | | yes[b] | 1.51 | 0.00 |
| | chr1:119929089 | *NOTCH2* | missense | c.3779G>A | p.Arg1260His | 3782 | 2002 | 1780 | 47.07 | | | | |
| EC_38-T2 | chr17:7675064 | *TP53* | nonsense | c.548C>A | p.S183* | 4853 | 3076 | 1777 | 36.62 | Ion S5 | yes[b] | 1.22 | 0.88 |
| | chr17:7676118 | *TP53* | frameshift deletion | c.250delG | p.Ala84fs | 6235 | 4490 | 1745 | 27.99 | | yes[b] | 1.51 | 0.00 |
| EC_39 | chr17:7577120 | *TP53* | missense | c.817C>T | p.Arg273Cys | 411 | 294 | 117 | 28.47 | Ion Proton | yes | 0.76 | 0.00 |
| | chr17:7578469 | *TP53* | missense | c.461G>T | p.Gly154Val | 135 | 115 | 20 | 14.81 | | yes[b] | 1.24 | 0.00 |
| EC_42 | chr9:136505109 | *NOTCH1* | splicesite | c.4587-5delC | | 5100 | 4567 | 533 | 10.45 | Ion S5 | NV[c] | | |
| EC_43 | chr17:7673740 | *TP53* | nonsense | c.880G>T | p.Glu294* | 3099 | 2198 | 901 | 29.07 | Ion S5 | yes | 0.09 | 0.00 |
| | chr4:153247289 | *FBXW7* | missense | c.1513C>G | p.Arg505Gly | 2000 | 1364 | 636 | 31.80 | | | | |
| EC_44 | chr8:38287382 | *FGFR1* | missense | c.275A>T | p.Asp92Val | 1967 | 1244 | 723 | 36.76 | Ion Proton | yes | 27.49 | 37.80 |
| | chr17:7577094 | *TP53* | missense | c.844C>T | p.Arg282Trp | 1794 | 400 | 1394 | 77.62 | | yes | 0.57 | 0.23 |
| | chr17:7579326 | *TP53* | frameshift deletion | c.353_360delCAGCCAAG | p.Thr118fs | 1987 | 1572 | 415 | 20.88 | | yes | 17.61 | 0.00 |
| EC_45 | chr17:80790226 | *ZNF750* | nonsense | c.104_105insA | p.Asn35Lys | 1986 | 1383 | 603 | 30.36 | Ion Proton | yes | 11.36 | 0.00 |
| | chr19:10597494 | *KEAP1* | missense | c.1709G>T | p.Gly570Val | 2000 | 1761 | 239 | 11.95 | | | | |
| EC_46 | chr9:136544021 | *NOTCH1* | splicesite | c.140+3G>T | | 2128 | 440 | 1688 | 79.32 | Ion S5 | yes[b] | 3.21 | 0.86 |
| | chr17:7675088 | *TP53* | missense | c.524G>A | p.Arg175His | 2167 | 1175 | 992 | 45.78 | | yes | 1.78 | 0.39 |
| EC_51 | chr17:7577518 | *TP53* | missense | c.763A>T | p.Ile255Phe | 1985 | 1822 | 162 | 8.20 | Ion Proton | yes[b] | 0.27 | 0.00 |
| | chr2:178098965 | *NFE2L2* | missense | c.80A>T | p.Asp27Val | 177 | 166 | 11 | 6.21 | | yes[b] | 0.00 | 0.00 |
| EC_52 | chr7:151960114 | *KMT2C* | missense | c.1286G>A | p.Gly429Asp | 2000 | 1506 | 494 | 24.70 | Ion Proton | | | |
| | chr17:7578551 | *TP53* | missense | c.379T>C | p.Ser127Pro | 128 | 111 | 17 | 13.28 | | yes[b] | 3.40 | 0.25 |
| | chr2:178098804 | *NFE2L2* | missense | c.241G>A | p.Gly81Ser | 2000 | 1622 | 378 | 18.90 | | yes | 0.65 | 0.06 |
| EC_54 | chr3:30713361 | *TGFBR2* | missense | c.761C>T | p.Ser254Phe | 417 | 337 | 80 | 19.18 | Ion Proton | | | |
| | chr17:7578190 | *TP53* | missense | c.659A>G | p.Tyr220Cys | 1993 | 1566 | 427 | 21.40 | | yes | 0.82 | 0.07 |
| | chr9:139413086 | *NOTCH1* | missense | c.1056C>A | p.Asp352Glu | 1511 | 1401 | 110 | 7.28 | | yes[b] | 0.00 | 0.00 |
| EC_55 | chr19:10602322 | *KEAP1* | missense | c.1256G>A | p.Gly419Glu | 1317 | 1209 | 108 | 8.20 | Ion Proton | NV[c] | | |

| | | | | | | | | | | | | | |
|---|---|---|---|---|---|---|---|---|---|---|---|---|---|
| | chr12:49444990 | *KMT2D* | missense | c.2476C>T | p.Pro826Ser | 367 | 330 | 37 | 10.08 | | | | |
| EC_56 | chr9:139401888 | *NOTCH1* | missense | c.3512G>T | p.Cys1171Phe | 1998 | 1092 | 906 | 45.35 | Ion Proton | | | |
| | chr17:7577537 | *TP53* | missense | c.743_744delGGinsAA | p.Arg248Gln | 1987 | 1000 | 987 | 49.60 | | yes | 0.28 | 0.06 |
| | chrX:44949063 | *KDM6A* | nonsense | c.3624T>A | p.Tyr1208Ter | 337 | 141 | 196 | 58.16 | | yes | 0.19 | 0.21 |
| EC_57 | chr14:23445861 | *AJUBA* | missense | c.1169A>G | p.Asp390Gly | 409 | 315 | 94 | 22.98 | Ion Proton | | | |
| | chr17:7577070 | *TP53* | nonsense | c.859G>T | p.Glu287Ter | 509 | 369 | 140 | 27.50 | | yes | 5.47 | 0.00 |
| | chr17:7577530 | *TP53* | missense | c.747G>T | p.Arg249Ser | 1968 | 1092 | 876 | 44.02 | | yes | 0.00 | 0.00 |
| EC_58 | chr10:89692905 | *PTEN* | missense | c.392C>A | p.Thr131Asn | 312 | 31 | 281 | 90.06 | Ion Proton | | | |
| | chr17:7579313 | *TP53* | missense | c.374C>T | p.Thr125Met | 1997 | 319 | 1678 | 83.90 | | yes | 15.65 | 3.37 |
| | chr17:80790218 | *ZNF750* | nonsense | c.113C>A | p.Ser38Ter | 1984 | 196 | 1788 | 90.12 | | yes | 28.37 | 6.50 |
| EC_59 | chr12:49050543 | *KMT2D* | frameshift insertion | c.3046_3047insT | p.Ile1015fs | 3489 | 2247 | 1242 | 35.60 | Ion S5 | | | |
| | chr16:3744932 | *CREBBP* | frameshift insertion | c.3944_3945insA | p.Thr1315fs | 1251 | 994 | 257 | 20.54 | | | | |
| | chr17:7674945 | *TP53* | nonsense | c.586C>T | p.Arg196* | 5788 | 2531 | 3257 | 56.27 | | yes | 0.63 | 0.06 |
| | chr22:41169571 | *EP300* | missense | c.4241T>C | p.Tyr1414Cys | 1624 | 1002 | 622 | 38.30 | | | | |
| EC_60 | chr5:151521372 | *FAT2* | frameshift deletion | c.11219_11220delCA | p.Thr3740fs | 3012 | 2288 | 724 | 24.04 | Ion S5 | | | |
| | chr17:7675211 | *TP53* | missense | c.401T>C | p.Phe134Ser | 118 | 39 | 79 | 66.95 | | yes[b] | 6.71 | 0.00 |
| EC_62 | chr4:186636719 | *FAT1* | missense | c.3838G>T | p.Asp1280Tyr | 878 | 631 | 247 | 28.13 | Ion S5 | | | |
| | chr17:7675994 | *TP53* | silent | c.375G>T | p.Thr125Thr | 2536 | 1606 | 930 | 36.67 | | yes[b] | 0.70 | 0.18 |
| EC_63 | chr17:7674928 | *TP53* | frameshift insertion | c.602_603insT | p.Leu201fs | 932 | 787 | 145 | 15.56 | Ion S5 | yes[b] | 2.54 | 0.00 |
| EC_64 | chr10:87933093 | *PTEN* | missense | c.853G>C | p.Leu285Val | 4032 | 3059 | 973 | 24.13 | Ion S5 | | | |
| | chr17:7673583 | *TP53* | frameshift insertion | c.944_945insGGCTC | p.Ser315fs | 2326 | 1818 | 508 | 21.84 | | yes | 0.16 | 0.11 |

[a] Plasma DNA was evaluated by dPCR
[b] Additional dPCR-evaluated mutations which not all of criterias were satisfied
[c] Designed dPCR probe could not be validated using tumor DNA.
[d] CtDNA VAFs analyzed by dPCR in plasma sample before or after initial cycle of chemotherapy

**Supplementary Table S4. Treatments and outcomes of the study patients**

| Patient ID | 1st line therapy | Number of cycles of 1st line therapy performed | Followed treatment with curative intent | Achieved to curative status | Relapse | Time from initiation of treatment to relapse (months) | Relapse site | Outcome |
|---|---|---|---|---|---|---|---|---|
| EC_2 | DCF | 6 | surgery | yes | yes | 57.5 | esophagus | alive with cancer |
| EC_3 | DCF | 2 | | no | | | | cancer death |
| EC_4 | DCF | 2 | CRT | no | | | | cancer death |
| EC_6 | CF | 2 | surgery | yes | yes | 59.2 | lymph node | alive with cancer |
| EC_7 | DCF | 1 | CRT | yes | no | | | alive |
| EC_8 | DCF | 3 | | no | | | | cancer death |
| EC_9 | DCF | 1 | | no | | | | cancer death |
| EC_10 | DCF | 6 | | no | | | | cancer death |
| EC_11 | DCF | 3 | | no | | | | cancer death |
| EC_13 | DCF | 4 | CRT | yes | yes | 20.9 | lymph node | alive |
| EC_14 | DCF | 2 | | no | | | | cancer death |
| EC_15 | DCF | 1 | | no | | | | cancer death |
| EC_16 | DCF | 1 | surgery / CRT | yes | yes | 20.9 | liver | cancer death |
| EC_18 | DCF | 2 | CRT | no | | | | alive with cancer |
| EC_19 | DCF | 2 | | no | | | | cancer death |
| EC_20 | DCF | 3 | surgery | yes | no | | | alive |
| EC_22 | DCF | 3 | surgery | yes | no | | | alive |
| EC_24 | DCF | 3 | CRT | no | | | | cancer death |
| EC_26 | DCF | 3 | CRT | no | | | | cancer death |
| EC_29 | DCF | 1 | CRT | no | | | | cancer death |
| EC_32 | DCF | 4 | surgery | yes | no | | | alive |
| EC_35 | DCF | 4 | CRT | no | | | | cancer death |
| EC_36 | CF | 2 | surgery | yes | yes | 32.1 | lymph node | alive with cancer |
| EC_38 | IC | 4 | surgery | no | | | | cancer death |
| EC_39 | DCF | 2 | surgery | yes | no | | | alive |
| EC_42 | DCF | 1 | | no | | | | cancer death |

| | | | | | | | | |
|---|---|---|---|---|---|---|---|---|
| EC_43 | DCF | 4 | CRT | yes | no | | | alive |
| EC_44 | DCF | 2 | CRT | no | | | | cancer death |
| EC_45 | DCF | 3 | CRT | yes | yes | 15.3 | lymph node | alive with cancer |
| EC_46 | DCF | 3 | CRT | no | | | | cancer death |
| EC_51 | DCF | 3 | CRT | no | | | | alive with cancer |
| EC_52 | DCF | 3 | CRT | no | | | | cancer death |
| EC_54 | DCF | 2 | surgery | yes | yes | 7.9 | liver, lymph node, bone(lumbar spine, rib), Myocardium | cancer death |
| EC_55 | DCF | 3 | surgery | yes | no | | | alive |
| EC_56 | DCF | 3 | surgery | no | | 4.7 | liver, lymph node | cancer death |
| EC_57 | DCF | 4 | surgery | yes | no | | | alive |
| EC_58 | CF | 6 | | no | | | | cancer death |
| EC_59 | CF | 2 | surgery | yes | yes | 18.4 | lymph node | alive with cancer |
| EC_60 | DCF | 3 | CRT | no | | | | non cancer death |
| EC_62 | DCF | 3 | surgery / CRT | yes | no | | | alive |
| EC_63 | DCF | 2 | CRT | no | | | | cancer death |
| EC_64 | DCF | 3 | surgery / CRT | yes | yes | 5.4 | lymph node | alive with cancer |

CRT, chemoradiotherapy; IC, irinotecan/cisplatin.

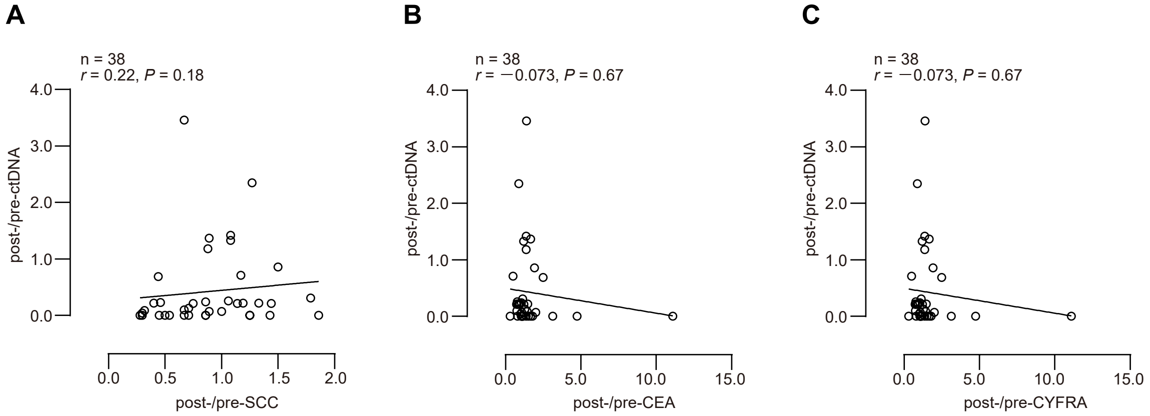


**Supplementary Fig. S1. Correlation between early change of ctDNA and conventional tumor markers.**
Correlation between **(A)** post-/pre-ctDNA and post-/pre-SCC, **(B)** post-/pre-ctDNA and post-/pre-CEA, and **(C)** post-/pre-ctDNA and post-/pre-SCC **(C)**, (Spearman’ s rank correlation coefficient).

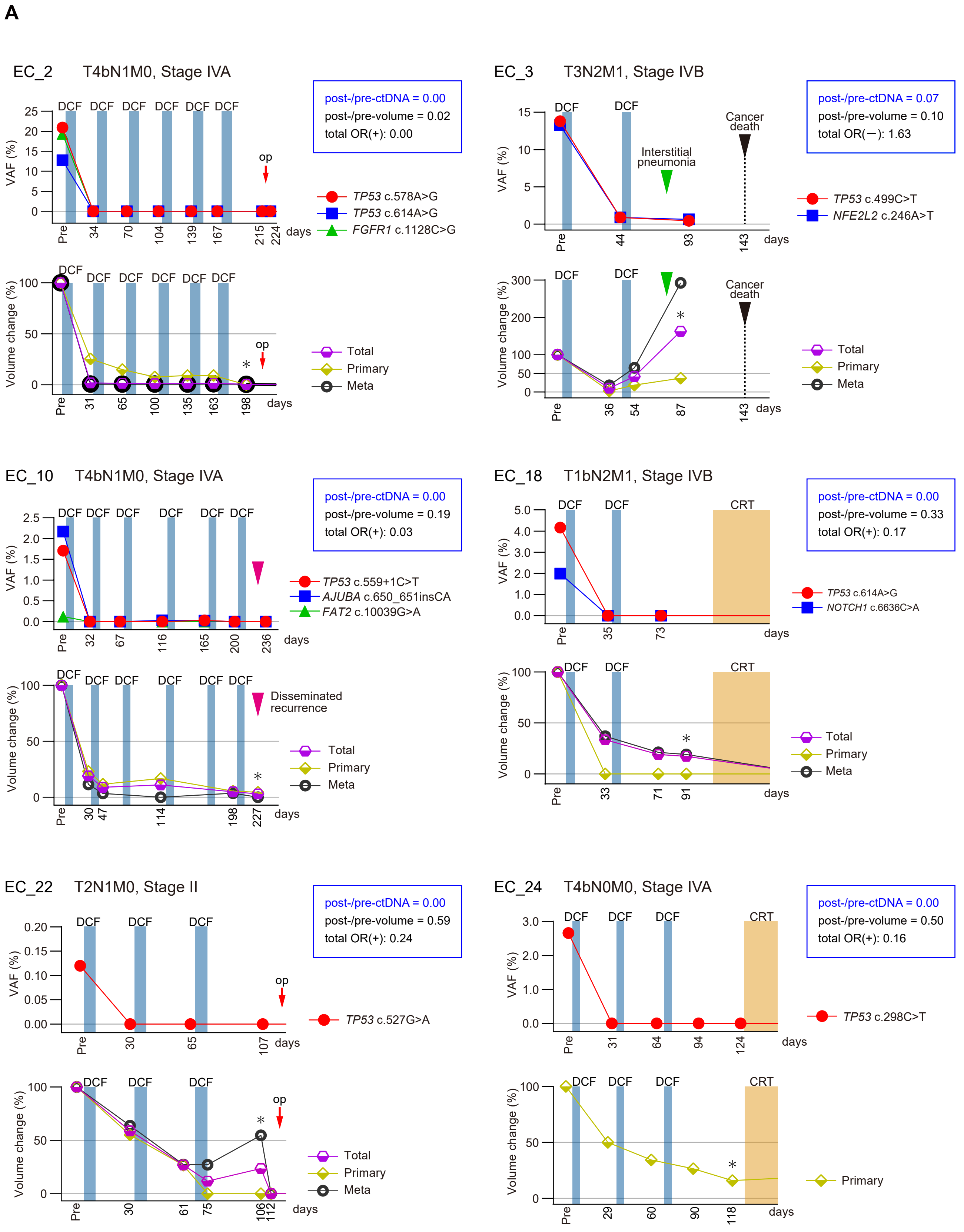
A
EC_2 T4bN1M0, Stage IVA
post-/pre-ctDNA = 0.00
post-/pre-volume = 0.02
total OR(+): 0.00
TP53 c.578A>G
TP53 c.614A>G
FGFR1 c.1128C>G
EC_3 T3N2M1, Stage IVB
post-/pre-ctDNA = 0.07
post-/pre-volume = 0.10
total OR(−): 1.63
Interstitial pneumonia
Cancer death
TP53 c.499C>T
NFE2L2 c.246A>T
EC_10 T4bN1M0, Stage IVA
post-/pre-ctDNA = 0.00
post-/pre-volume = 0.19
total OR(+): 0.03
TP53 c.559+1C>T
AJUBA c.650_651insCA
FAT2 c.10039G>A
Disseminated recurrence
EC_18 T1bN2M1, Stage IVB
post-/pre-ctDNA = 0.00
post-/pre-volume = 0.33
total OR(+): 0.17
TP53 c.614A>G
NOTCH1 c.6636C>A
EC_22 T2N1M0, Stage II
post-/pre-ctDNA = 0.00
post-/pre-volume = 0.59
total OR(+): 0.24
TP53 c.527G>A
EC_24 T4bN0M0, Stage IVA
post-/pre-ctDNA = 0.00
post-/pre-volume = 0.50
total OR(+): 0.16
TP53 c.298C>T
DCF
CRT
op
VAF (%)
Volume change (%)
days
Pre
Total
Primary
Meta

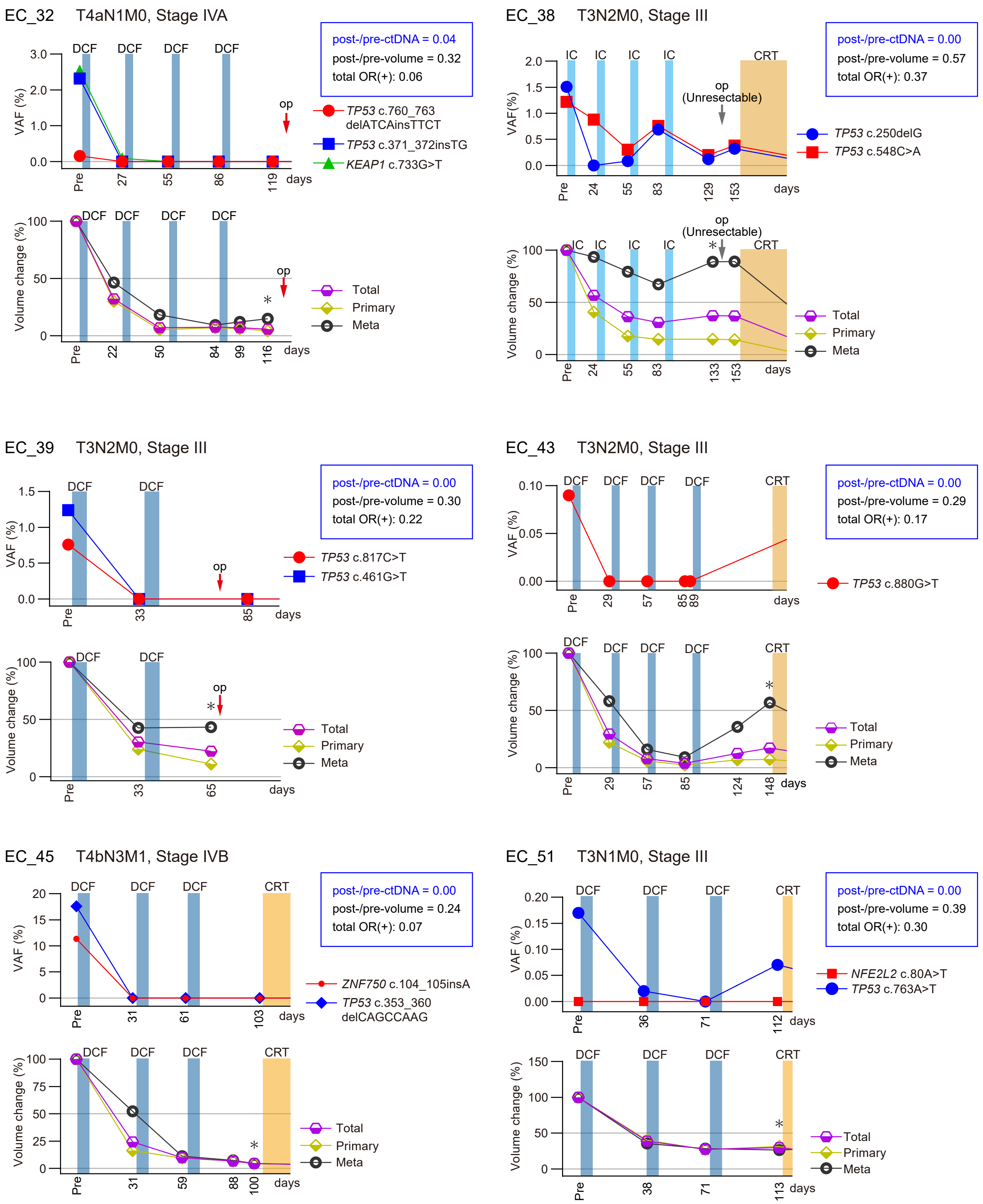

EC_32 T4aN1M0, Stage IVA
post-/pre-ctDNA = 0.04
post-/pre-volume = 0.32
total OR(+): 0.06
TP53 c.760_763 delATCAinsTTCT
TP53 c.371_372insTG
KEAP1 c.733G>T
VAF (%)
Volume change (%)
DCF
op
days
Total
Primary
Meta
EC_38 T3N2M0, Stage III
post-/pre-ctDNA = 0.00
post-/pre-volume = 0.57
total OR(+): 0.37
TP53 c.250delG
TP53 c.548C>A
IC
CRT
op (Unresectable)
EC_39 T3N2M0, Stage III
post-/pre-ctDNA = 0.00
post-/pre-volume = 0.30
total OR(+): 0.22
TP53 c.817C>T
TP53 c.461G>T
EC_43 T3N2M0, Stage III
post-/pre-ctDNA = 0.00
post-/pre-volume = 0.29
total OR(+): 0.17
TP53 c.880G>T
EC_45 T4bN3M1, Stage IVB
post-/pre-ctDNA = 0.00
post-/pre-volume = 0.24
total OR(+): 0.07
ZNF750 c.104_105insA
TP53 c.353_360 delCAGCCAAG
EC_51 T3N1M0, Stage III
post-/pre-ctDNA = 0.00
post-/pre-volume = 0.39
total OR(+): 0.30
NFE2L2 c.80A>T
TP53 c.763A>T

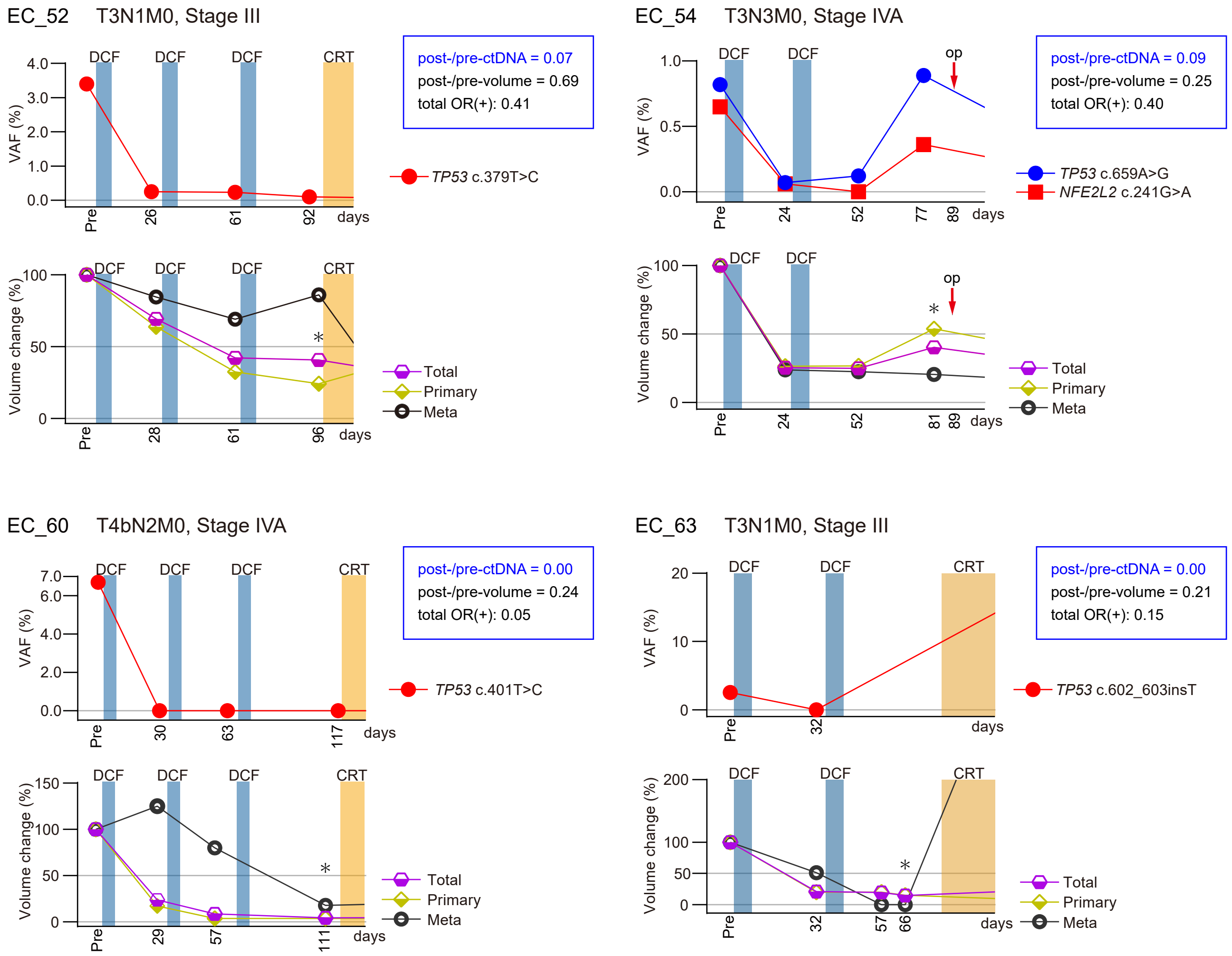
EC_52 T3N1M0, Stage III
DCF
DCF
DCF
CRT
VAF (%)
4.0
3.0
2.0
1.0
0.0
Pre
26
61
92
days
post-/pre-ctDNA = 0.07
post-/pre-volume = 0.69
total OR(+): 0.41
TP53 c.379T>C
Volume change (%)
100
50
0
Pre
28
61
96
days
Total
Primary
Meta
EC_54 T3N3M0, Stage IVA
DCF
DCF
op
VAF (%)
1.0
0.5
0.0
Pre
24
52
77
89
days
post-/pre-ctDNA = 0.09
post-/pre-volume = 0.25
total OR(+): 0.40
TP53 c.659A>G
NFE2L2 c.241G>A
Volume change (%)
100
50
0
Pre
24
52
81
89
days
Total
Primary
Meta
EC_60 T4bN2M0, Stage IVA
DCF
DCF
DCF
CRT
VAF (%)
7.0
6.0
4.0
2.0
0.0
Pre
30
63
117
days
post-/pre-ctDNA = 0.00
post-/pre-volume = 0.24
total OR(+): 0.05
TP53 c.401T>C
Volume change (%)
150
100
50
0
Pre
29
57
111
days
Total
Primary
Meta
EC_63 T3N1M0, Stage III
DCF
DCF
CRT
VAF (%)
20
10
0
Pre
32
days
post-/pre-ctDNA = 0.00
post-/pre-volume = 0.21
total OR(+): 0.15
TP53 c.602_603insT
Volume change (%)
200
100
50
0
Pre
32
57
66
days
Total
Primary
Meta

**B**

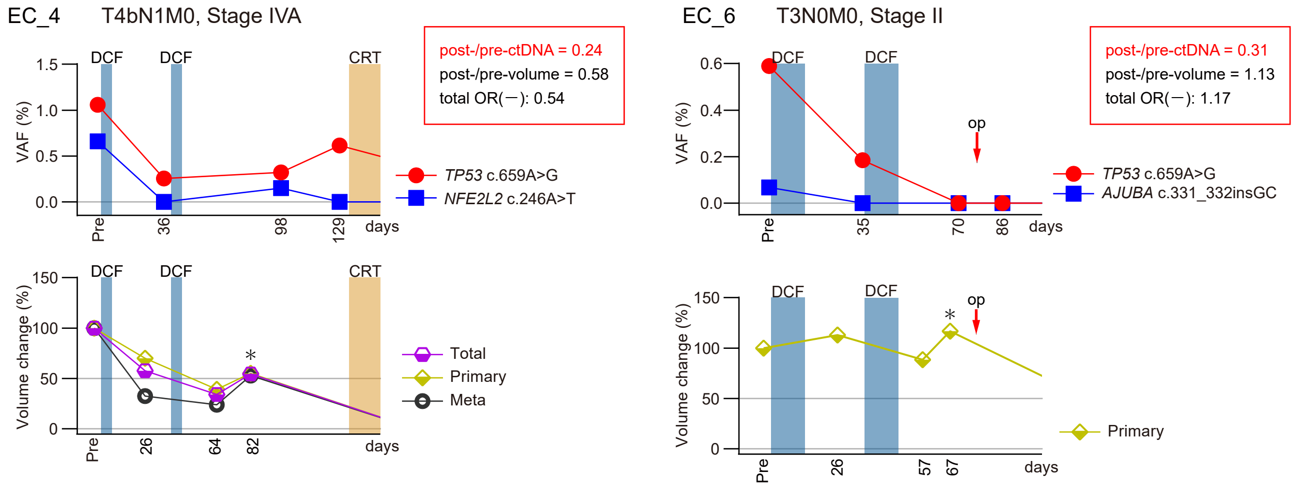
EC_4 T4bN1M0, Stage IVA
DCF
DCF
CRT
VAF (%)
1.5
1.0
0.5
0.0
Pre
36
98
129
days
post-/pre-ctDNA = 0.24
post-/pre-volume = 0.58
total OR(−): 0.54
TP53 c.659A>G
NFE2L2 c.246A>T
Volume change (%)
150
100
50
0
Pre
26
64
82
days
Total
Primary
Meta
EC_6 T3N0M0, Stage II
DCF
DCF
op
VAF (%)
0.6
0.4
0.2
0.0
Pre
35
70
86
days
post-/pre-ctDNA = 0.31
post-/pre-volume = 1.13
total OR(−): 1.17
TP53 c.659A>G
AJUBA c.331_332insGC
Volume change (%)
150
100
50
0
Pre
26
57
67
days
Primary

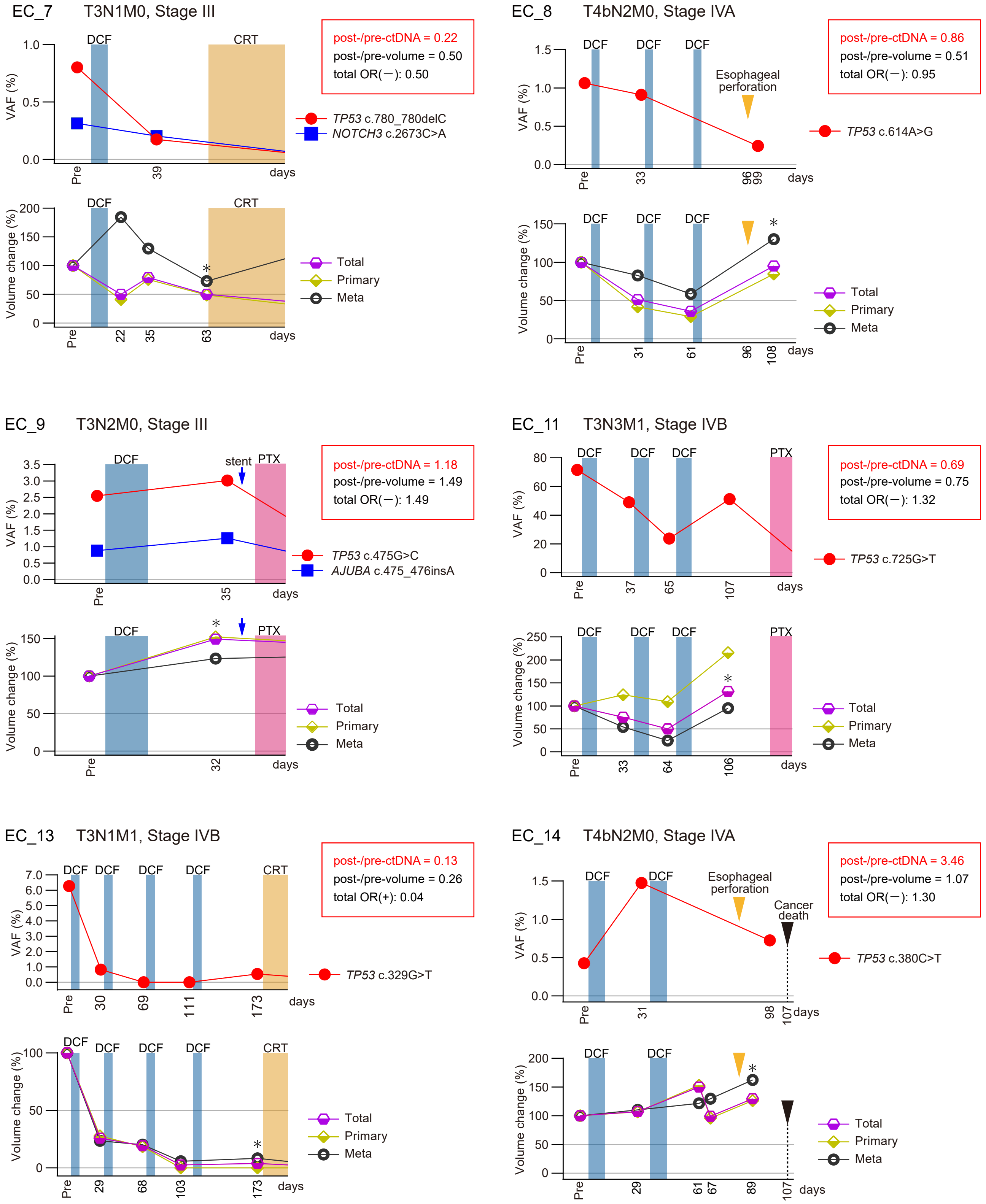

EC_7 T3N1M0, Stage III
post-/pre-ctDNA = 0.22
post-/pre-volume = 0.50
total OR(−): 0.50
TP53 c.780_780delC
NOTCH3 c.2673C>A
DCF
CRT
VAF (%)
Volume change (%)
Total
Primary
Meta
days
EC_8 T4bN2M0, Stage IVA
post-/pre-ctDNA = 0.86
post-/pre-volume = 0.51
total OR(−): 0.95
Esophageal perforation
TP53 c.614A>G
EC_9 T3N2M0, Stage III
post-/pre-ctDNA = 1.18
post-/pre-volume = 1.49
total OR(−): 1.49
stent
PTX
TP53 c.475G>C
AJUBA c.475_476insA
EC_11 T3N3M1, Stage IVB
post-/pre-ctDNA = 0.69
post-/pre-volume = 0.75
total OR(−): 1.32
TP53 c.725G>T
EC_13 T3N1M1, Stage IVB
post-/pre-ctDNA = 0.13
post-/pre-volume = 0.26
total OR(+): 0.04
TP53 c.329G>T
EC_14 T4bN2M0, Stage IVA
post-/pre-ctDNA = 3.46
post-/pre-volume = 1.07
total OR(−): 1.30
Esophageal perforation
Cancer death
TP53 c.380C>T

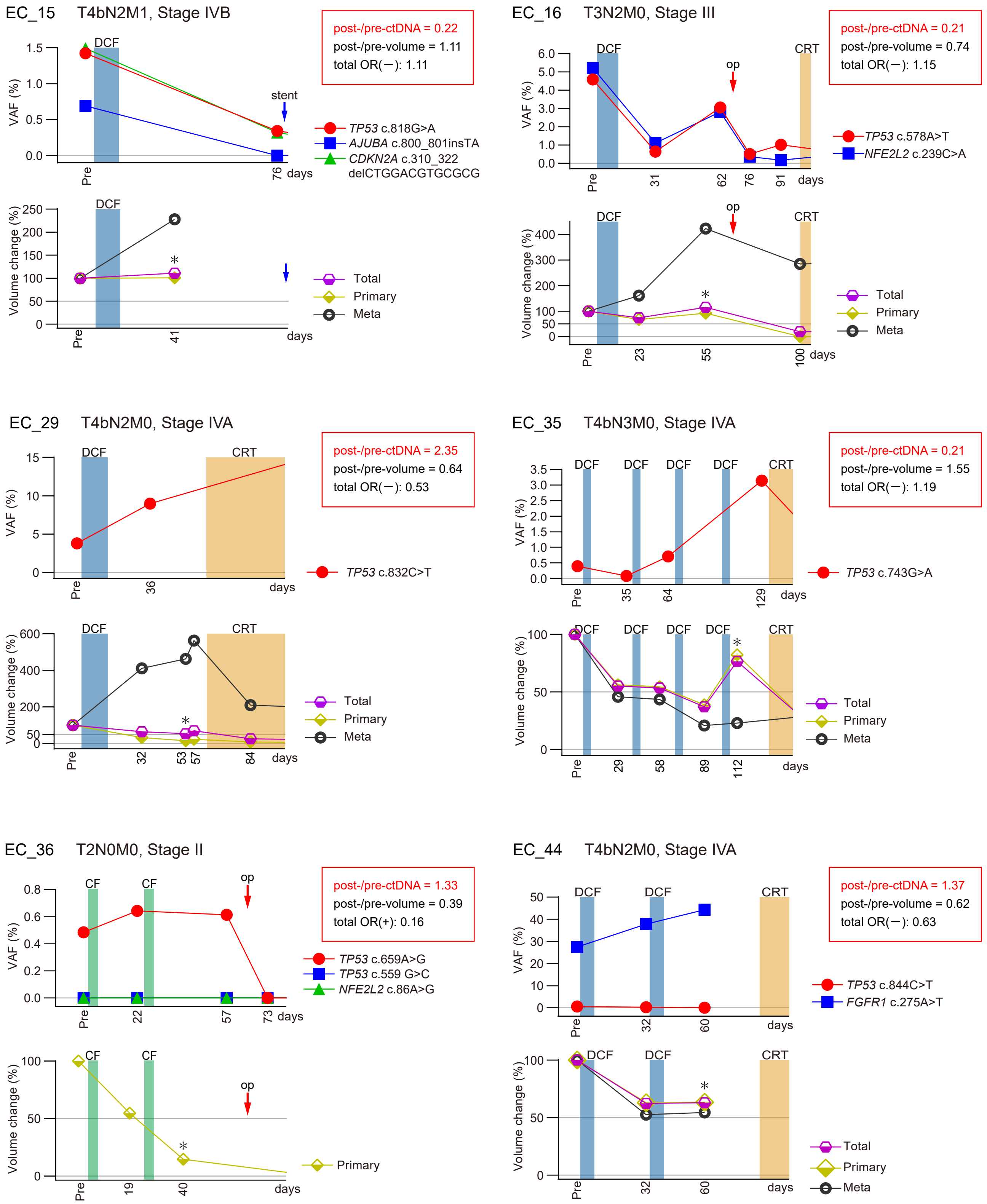

EC_15 T4bN2M1, Stage IVB
post-/pre-ctDNA = 0.22
post-/pre-volume = 1.11
total OR(−): 1.11
VAF (%)
DCF
stent
Pre
76
days
TP53 c.818G>A
AJUBA c.800_801insTA
CDKN2A c.310_322 delCTGGACGTGCGCG
Volume change (%)
*
41
Total
Primary
Meta
EC_16 T3N2M0, Stage III
post-/pre-ctDNA = 0.21
post-/pre-volume = 0.74
total OR(−): 1.15
DCF
op
CRT
31
62
76
91
TP53 c.578A>T
NFE2L2 c.239C>A
23
55
100
EC_29 T4bN2M0, Stage IVA
post-/pre-ctDNA = 2.35
post-/pre-volume = 0.64
total OR(−): 0.53
36
TP53 c.832C>T
32
53
57
84
EC_35 T4bN3M0, Stage IVA
post-/pre-ctDNA = 0.21
post-/pre-volume = 1.55
total OR(−): 1.19
35
64
129
TP53 c.743G>A
29
58
89
112
EC_36 T2N0M0, Stage II
post-/pre-ctDNA = 1.33
post-/pre-volume = 0.39
total OR(+): 0.16
CF
22
57
73
TP53 c.659A>G
TP53 c.559 G>C
NFE2L2 c.86A>G
19
40
EC_44 T4bN2M0, Stage IVA
post-/pre-ctDNA = 1.37
post-/pre-volume = 0.62
total OR(−): 0.63
32
60
TP53 c.844C>T
FGFR1 c.275A>T

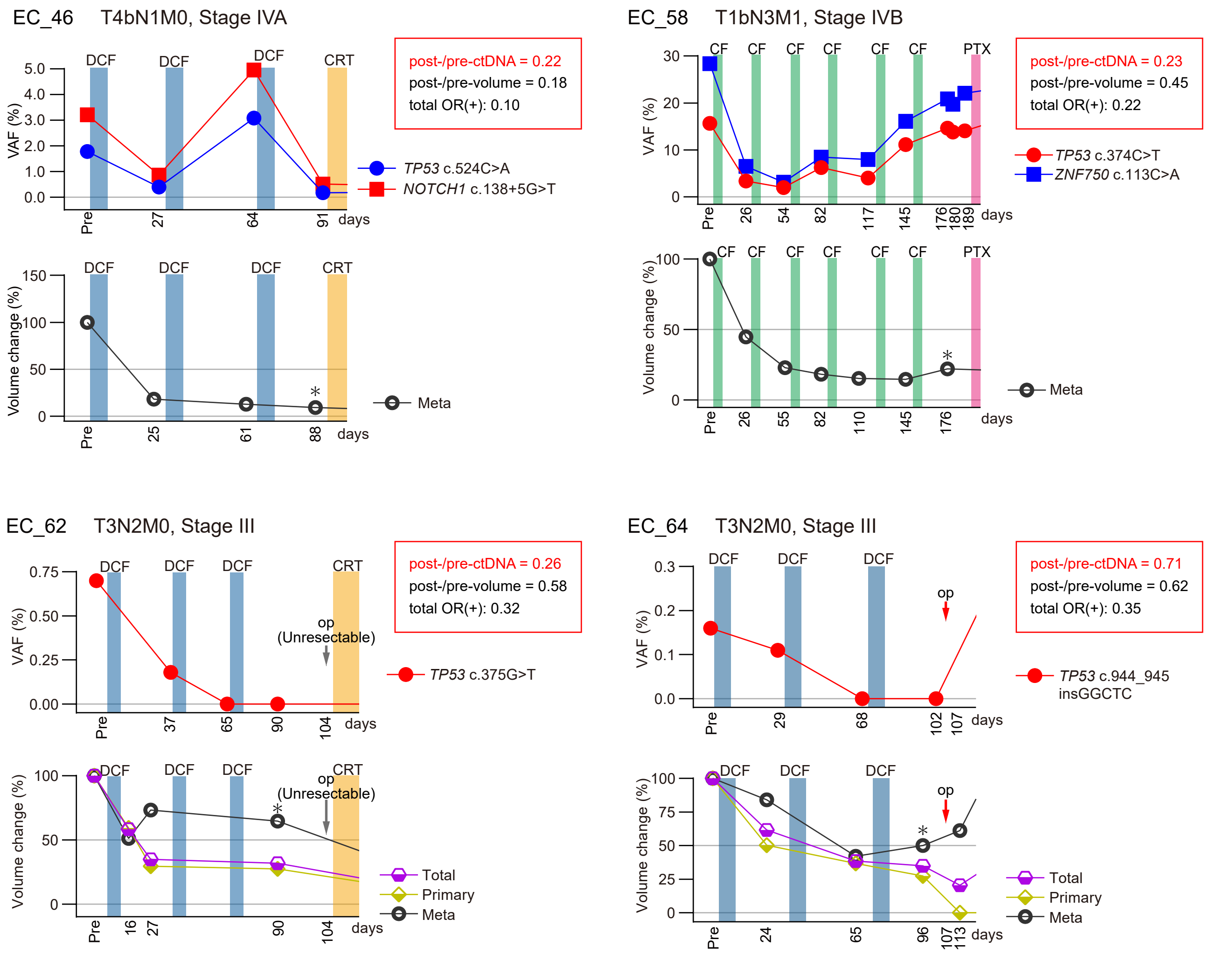


**Supplementary Fig. S2. Dynamics of ctDNA and tumor volume during first-line chemotherapy in the study patients.**

**(A)** Patients with low post-/pre-ctDNA. **(B)** Patients with high post-/pre-ctDNA. Patients with levels of post-/pre-ctDNA below and above the cutoff value of 0.13 were assigned to low and high post-/pre-ctDNA groups, respectively. The asterisk (*) indicates the timepoint of the CT in which the total OR was assessed. CRT; chemotherapy; IC, irinotecan/cisplatin; op, operation; PTX, paclitaxel.

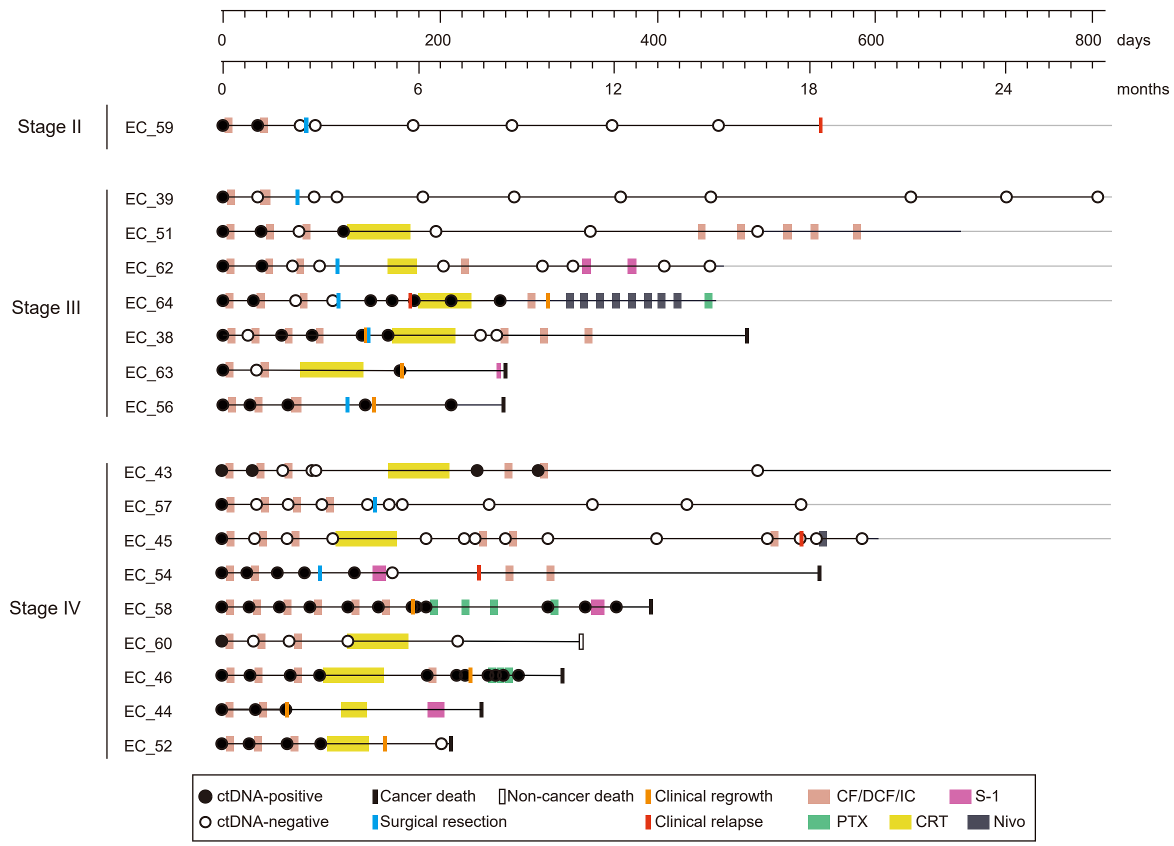


**Supplementary Fig. S3. Longitudinal ctDNA monitoring and clinical information.**
The status of ctDNA and clinical information are schematized on the horizontal lines. CRT, chemotherapy; IC, irinotecan/cisplatin; Nivo, nivolumab; PTX, paclitaxel.
